# Analysis of respiratory and systemic immune responses in COVID-19 reveals mechanisms of disease pathogenesis

**DOI:** 10.1101/2020.10.15.20208041

**Authors:** Peter A. Szabo, Pranay Dogra, Joshua I. Gray, Steven B. Wells, Thomas J. Connors, Stuart P. Weisberg, Izabela Krupska, Rei Matsumoto, Maya M.L. Poon, Emma Idzikowski, Sinead E. Morris, Chloé Pasin, Andrew J. Yates, Amy Ku, Michael Chait, Julia Davis-Porada, Jing Zhou, Matthew Steinle, Sean Mackay, Anjali Saqi, Matthew Baldwin, Peter A. Sims, Donna L. Farber

## Abstract

Immune responses to respiratory viruses like SARS-CoV-2 originate and function in the lung, yet assessments of human immunity are often limited to blood. Here, we conducted longitudinal, high-dimensional profiling of paired airway and blood samples from patients with severe COVID-19, revealing immune processes in the respiratory tract linked to disease pathogenesis. Survival from severe disease was associated with increased CD4^+^T cells and decreased monocyte/macrophage frequencies in the airway, but not in blood. Airway T cells and macrophages exhibited tissue-resident phenotypes and activation signatures, including high level expression and secretion of monocyte chemoattractants CCL2 and CCL3 by airway macrophages. By contrast, monocytes in blood expressed the CCL2-receptor CCR2 and aberrant CD163^+^ and immature phenotypes. Extensive accumulation of CD163^+^monocyte/macrophages within alveolar spaces in COVID-19 lung autopsies suggested recruitment from circulation. Our findings provide evidence that COVID-19 pathogenesis is driven by respiratory immunity, and rationale for site-specific treatment and prevention strategies.

## INTRODUCTION

The novel respiratory virus SARS-CoV-2 has resulted in devastating impacts to the world’s population, both as a result of morbidity and mortality caused by COVID-19, as well as the life-altering measures implemented to mitigate spread. While the majority of infected individuals (>90%) develop a self-limiting disease and recover, approximately 5-10% of individuals develop severe respiratory disease marked by lung infiltrates and reduced oxygen saturation, which can progress to acute respiratory distress syndrome (ARDS), multi-organ failure, and death (Wu and McGoogan, 2020). Risk factors for severe COVID-19 include older age and co-morbidities like obesity and diabetes, although younger and previously healthy individuals can also be susceptible (Cummings et al., 2020; Davies et al., 2020). For individuals who recover from self-limiting illness, the immune system acts in a coordinated fashion to clear the virus and establish virus-specific immunity (Moderbacher et al., 2020). However, the role of the immune response in the pathogenesis of severe COVID-19 remains unclear, and understanding this phenomenon is urgently required to develop new treatment and prevention strategies.

A key aspect of the immune response to respiratory virus infection is the activation and mobilization of immune cells to the lung for viral clearance. Innate immune responses are initiated within infected lung epithelial cells and local immune cells, including tissue resident alveolar macrophages and infiltrating monocytes and granulocytes (Yoo et al., 2013). The initial production of pro-inflammatory cytokines in the lung can precipitate cytokine storms in severe respiratory infections (Teijaro et al., 2011b). Adaptive immune responses are also mobilized in the lung; antigen-loaded dendritic cells migrate from the infected lung to the draining lymph node where they prime CD4^+^ and CD8^+^T cells. The resultant effector T cells traffic to the lung to mediate clearance of infected cells *in situ* (Yoo et al., 2013). A proportion of these lung effector T cells develop into tissue resident memory T cells (TRM), which are retained in the lung and can mediate rapid protective responses upon viral challenge in mouse models (Teijaro et al., 2011a; Turner et al., 2014; Turner and Farber, 2014; Wu et al., 2014). TRM in mice and humans are phenotypically and transcriptionally distinct from circulating effector-memory (TEM) cells (Kumar et al., 2017; Mackay et al., 2016; Masopust and Soerens, 2019). In adult lungs, TRM are the predominant T cell subset and persist in stable frequencies throughout life (Kumar et al., 2018), suggesting a crucial role in protection to respiratory pathogens. Moreover, CD4^+^TRM-like cells in the airway are required for protection against SARS-CoV-1 in mice (Zhao et al., 2016). At present, we lack information on the role of resident immune cells, including alveolar macrophages and lung TRM in protection against SARS-CoV-2 infection, and their function in the pathogenesis of severe COVID-19.

Studies of the immune response to SARS-CoV-2 have examined innate and adaptive immune cells, as well as soluble mediators in blood and plasma of infected individuals, revealing elevated levels of pro-inflammatory cytokines (Hadjadj et al., 2020; Laing et al., 2020) and robust virus-specific adaptive immune responses. Virus-specific CD4^+^ and CD8^+^T cells are found in most infected individuals with varying disease severities and persist following recovery (Grifoni et al., 2020; Thieme et al., 2020; Weiskopf et al., 2020). Antibodies specific for different viral proteins, including anti-Spike (S) protein-specific neutralizing antibodies (Long et al., 2020; Ni et al., 2020), also persist after resolution. How these systemic immune responses relate to innate and adaptive immunity in the respiratory tract is unclear and difficult to assess.

Here, we present in-depth, high-dimensional profiling of innate and adaptive immune cells and their functional responses in paired airway and blood samples obtained longitudinally from 15 patients with severe COVID-19, along with control airway and blood samples. We identified robust innate and adaptive immune responses in the airway, which were distinct from blood in cellular composition, function and transcriptional profile. Notably, increased frequencies CD4^+^T cells and decreased frequencies monocytes/macrophages in airways were associated with survival and younger age, suggesting key roles for these cells at the infection site. COVID-19 airways contained activated TRM, high frequencies of inflammatory tissue monocytes/macrophages, and supranormal levels of the monocyte chemoattractant cytokines CCL2, CCL3, and CCL4 — all lacking in blood, which contained predominant populations of immature monocytes. Excessive macrophage/monocyte content in COVID-19 lung autopsies compared to control lungs provide evidence for dynamic monocyte recruitment to the respiratory tract. Our results reveal compartmentalization of innate and adaptive immune responses in the respiratory tract of COVID-19, which drives peripheral immune cell infiltration and disease pathogenesis.

## RESULTS

### Obtaining paired airway and blood samples from severe COVID-19 patients

During the height of the pandemic in New York City, between April and June 2020, we enrolled patients from adult and pediatric intensive care units at New York Presbyterian hospital with severe COVID-19 (confirmed by positive SARS-CoV-2 PCR). Enrolled patients required mechanical ventilator support enabling us to obtain paired airway and blood samples longitudinally for up to 10 days during their hospitalization (average 6-7 sample days per patient). Sampling for each patient began within 24-36 hours of intubation. Patients represented a broad age range (14-84 yrs) and 8/15 (53%) died during enrollment or soon after (Table S1). Enrolled COVID-19 patients exhibited similar clinical severity measures regardless of outcome; the extent of ARDS, inflammation, neutrophil levels and comorbidities were similar between deceased patients and those who survived, while median age differed significantly (72yrs, deceased; 39yrs, survived). All patients developed robust SARS-CoV-2-specific neutralizing antibodies as measured in plasma (Weisberg et al., 2020).

Airway samples were obtained using a saline wash of the endotracheal tube performed daily as part of clinical care, which we have previously shown contain respiratory immune cell populations (Connors et al., 2018; Connors et al., 2016). A total of 141 paired blood and airway cell preparations were analyzed by high-dimensional spectral flow cytometry and successive samples from four patients were profiled by scRNAseq (Figure 1A, Table S2, S3). Airway supernatants and blood plasma from early and late time points were also assayed for cytokines and chemokines (Figure 1A, Table S2).

**Figure 1.**
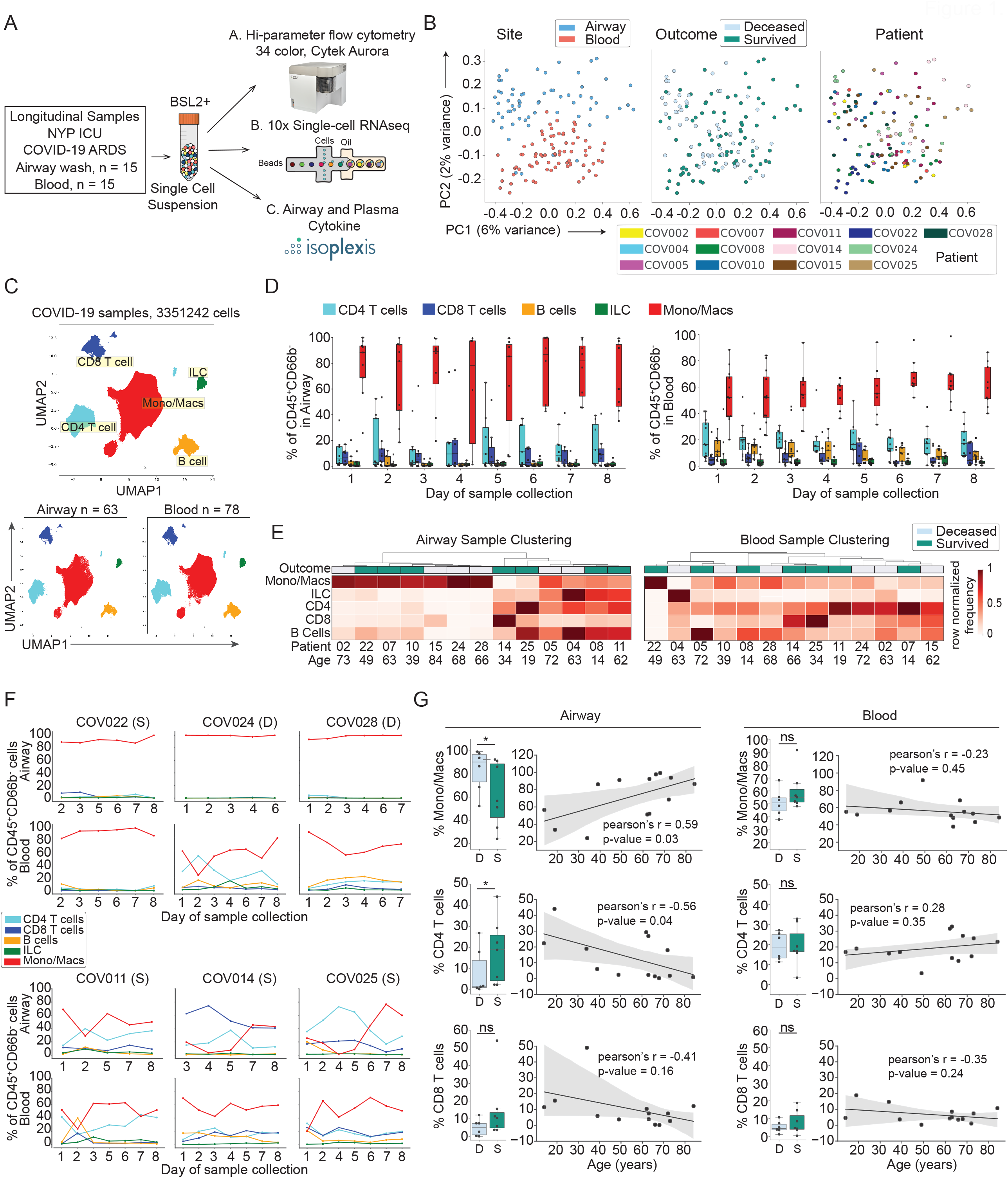
Distinct immune cell composition in airways compared to blood is associated with outcome and age. (A) Schematic diagram showing assays performed on COVID-19 patient airway and blood samples for this study. (B) Principal component analysis (PCA) of all COVID-19 samples based on mean marker expression colored by site (left), outcome (middle) and by donor (right). (C) UMAP embedding of flow cytometry results from all airway and blood samples combined colored by major cell lineage (top panel), and separated by tissue site (bottom two panels). (D) Immune cell composition over time in airways and blood. Box plots show the frequency of each major cell lineage of CD45^+^CD66B^-^ cells in airway (left) and blood (right) samples collected longitudinally for each sample day. Color of boxes corresponds to lineage and each dot is an individual patient sample. (E) Hierarchical clustering of airway (left) and blood (right) samples based on average lineage frequency across all time points for each donor-site group. Heatmaps are colored by row normalized value for each sample. (F) Line plots showing frequency of major lineages of total CD45^+^CD66B^-^ cells in airway (top row) and blood (bottom row) samples collected longitudinally for representative donors. (G) Association of monocyte/macrophage and T cell frequencies in airway (left) and blood (right) with outcome (deceased or survived) and correlation with age. Statistical significance was calculated using Mann-Whitney U-tests (box-plots) or Pearson correlations (scatter plots) and indicated by ***, p ≤ 0.001; **, p ≤ 0.01; *, p ≤ 0.05.

### Distinct immune cell composition in airway and blood of COVID-19 patients

Mononuclear cells from paired airway and blood samples were isolated by centrifugation through ficoll (see methods), stained using a 34 marker panel containing antibodies specific for major lineage determinants and markers for differentiation, tissue residence, activation, and function (see methods), and analyzed by spectral flow cytometry (gating strategy for mononuclear cells shown in Figure S1). Principal component analysis (PCA) of mean marker expression for each sample showed distinct clustering of airway and blood samples by site, but not by outcome or by patient (Figure 1B, Table S4). Computational analysis of flow cytometry data visualized by uniform manifold approximation and projection (UMAP) embedding (see methods) showed distinct separation of the major lineages into monocytes/macrophages, CD4^+^T cells, CD8^+^T cells, B cells, and innate lymphoid cells (predominantly NK cells) for all samples (Figure 1C, Figure S2A). Compiled data for each timepoint revealed distinct immune cell composition in airway compared to blood (Figure 1D). Airway samples had predominant frequencies of monocytes/macrophages (40-90%), lower T cell frequencies, and very low-to-negligible frequencies of B cells and ILCs, while blood contained higher lymphocyte frequencies with monocytes comprising ∼50% of all non-neutrophil leukocytes (Figure 1D). Similar immune cell compositions were confirmed by scRNAseq analysis of airway and blood from four individuals (Figure S2B). These results show distinct immune cell profiles in airway compared to blood across all patients and timepoints analyzed.

We investigated whether the airway or blood immune cell composition differentiated between patients or correlated with overall survival. Hierarchical clustering of aggregated samples from each individual revealed two major patterns of immune cell composition in the airways; one pattern showing a predominance of monocytes/macrophages, while the second pattern had higher frequencies of CD4^+^T cells, B cells, and ILCs compared to the first (Figure 1E left). However, neither pattern significantly correlated to outcome as patients who survived or succumbed were represented in both groups (Figure 1E, left). Hierarchical clustering of blood immune cell data revealed multiple clusters with different numbers of patients in each with no clear distinction in patterns or by patient outcome (Figure 1E, right). The longitudinal profiles of immune cell composition for individual patient samples also showed distinct composition in airways, which did not correspond to blood either in frequency or in changes over time (Figure 1F, Figure S3A). However, examination of specific lineages showed significant associations with outcome and correlation with age. Notably, there was a significant decrease in frequencies of airway monocytes/macrophages and an increase in airway CD4^+^T cells in patients who survived the disease versus those who succumbed (Figure 1G, left, Figure S3B), while the frequency of the corresponding blood immune cell subsets did not significantly differ between patients based on outcome, nor did they correlate with age (Figure 1G, right, Figure S3B). Accordingly, clustering the longitudinal patterns of cell type frequencies using K-means further suggests that airway immune cell trajectories are a better indicator of clinical outcome than their blood counterparts (Figure S3C). Together, these results show that airways exhibit an immune cell composition distinct from blood, and that the dynamics of airway T cells and monocyte/macrophages are significantly associated with outcome, suggesting key roles for these cell types in disease pathogenesis.

### Tissue resident memory T cells are the major T cell subset in airways

The subset composition and transcriptional profile of airway T cells in comparison to those in blood was further examined through high-dimensional, single cell approaches. Multiple markers of T cell differentiation were used to distinguish naïve and memory populations (CD45RA, CCR7, CD95, CD27), activation (HLA-DR, PD-1), functional subsets (FOXP3/CD25 for Tregs, CXCR5/PD-1 for Tfh-like, TCRGD for γδ T cells), specific states of senescence or terminal differentiation (CD57, KLRG1), and tissue residence (CD69, CD103). We used UMAP embedding to visualize expression of these multiple markers by airway and blood T cells, showing increased expression of CD69, CD103, PD-1, and HLA-DR in the airways and increased CCR7, CD45RA, and CD127 expression in the blood (Figure S4A and S4B). Phenograph clustering based on marker expression by CD4^+^ and CD8^+^T cells yielded 27 clusters, which were coalesced into 15 clusters denoting biological subsets or functional/activation states in airway and blood (Figure 2A, 2B).

**Figure 2.**
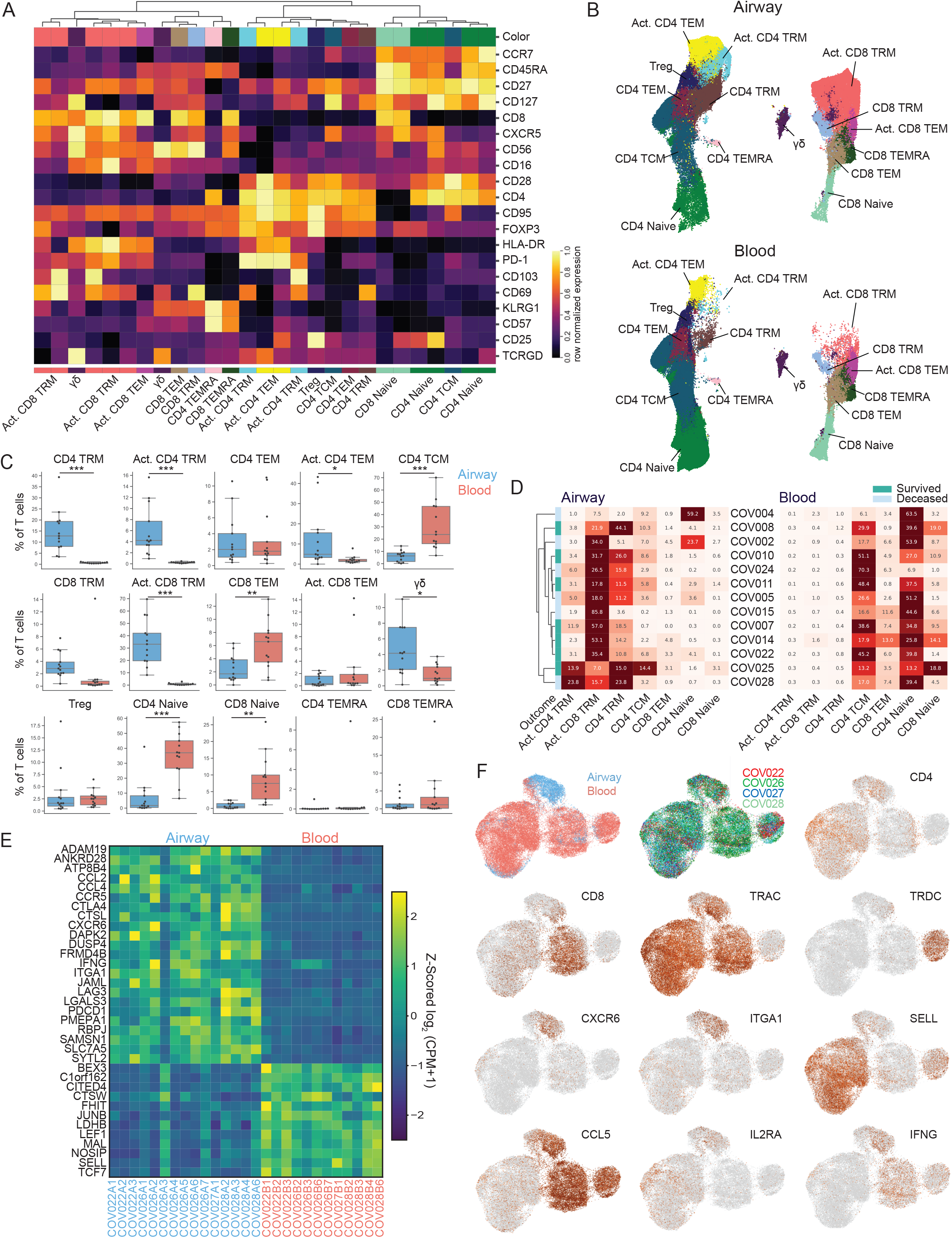
Airway T cells in COVID-19 are dominated by TRM and activated phenotypes. (A) Heatmap displaying expression of markers within phenograph-generated, hierarchical T cell clusters. The 27 phenograph clusters were collapsed into 15 definable cell subsets indicated at bottom. Heatmap data are colored by row normalized value for each sample. (B) UMAP embedding of 15 T cell subsets in the airway (upper) and blood (lower) with labels denoting the specific subset as defined in A. (C) T cell subset frequencies in airway compared to blood samples. Boxplots showing frequency of the indicated T cell subset for each patient (average of all time points collected) in the airway (blue) and blood (red). Statistical significance was calculated using a paired T-tests and indicated by ***, p ≤ 0.001; **, p ≤ 0.01; *, p ≤ 0.05. (D) Frequencies of the major T cell subsets in airway and blood shown for each individual patient and their outcome in airway (left) and blood (right) shown in a heat map. (only select subsets shown) (E) Heatmap showing major differentially expressed genes in airway compared to blood T cells by scRNA-seq from each individual patient and timepoint. Data are colored by row z-score for each sample. (F) Separate UMAP embeddings of gene expression by scRNA-seq from total T cells obtained from airway and blood of paired samples from four patients. UMAP shows airway (blue) and blood (red) origin of samples, patient, and indicated gene expression (based on Log2(CPM+1)).

There were significant qualitative and quantitative differences in T cell subset composition and activation state between airway and blood. In particular, airway contained CD4^+^ and CD8^+^TRM cells (CD69^+^CD103^+/-^) along with activated TRM subsets expressing elevated levels of HLA-DR and PD-1, and reduced levels of CD127 compared to non-activated TRM (Figure 2A, 2B). TRM cells, regardless of activation state, were largely confined to the airways and not significantly present in blood (Figure 2B, 2C), consistent with virus-responding T cells located at the site of infection. Innate-like γδ cells were also present in higher frequencies in airways compared to blood (Figure 2C). Circulating TEM cells (CD69^-^CD103^-^) were present in both sites, with non-activated CD8^+^TEM enriched in the blood (Figure 2B, 2C). Blood also contained higher frequencies of naïve CD4^+^ and CD8^+^T cells, CD4^+^TCM cells (Figure 2B, 2C). Between patients, there was variability in the proportions of the major subsets represented; most patients (9/13) had predominant CD8^+^TRM in airways, while 3/13 patients had higher frequencies of CD4^+^TRM in airways (Figure 2D). Together, these analyses indicate that both TRM and activated memory T cells (TRM and TEM) exhibit biased distribution in favor of the airways and not blood, and that the subset composition and activation states of blood T cells does reflect the dynamics in airways.

Consistent with the flow cytometry results, analysis of T cells by scRNA-seq revealed distinct transcriptional profiles expressed by airway compared to blood T cells. TRM signature genes *CXCR6* and *ITGA1* were uniquely expressed by airway T cells (Figure 2E, F, Table S5), consistent with previous scRNA-seq analysis of human TRM cells in lung and other sites (Snyder et al., 2019; Szabo et al., 2019). Naïve and TCM cells distinguished by *SELL* expression were highly enriched in blood, while *CCL5* expression indicating TEM cells were found in both sites (Figure 2F). Identification of the top differentially expressed genes between airway and blood revealed that T cells from the airway exhibit a gene signature associated with TRM and tissue T cells (Kumar et al., 2017; Szabo et al., 2019), including upregulated expression of *CXCR6, ITGA1, PDCD1, LGALS, LAG3*, and *RBPJ* compared to blood T cells (Figure 2E, F, Table S5). Airway T cells also showed upregulated expression of genes encoding key cytokines and chemokines, including *IFNG, CCL2*, and *CCL4* (Figure 2E, F), consistent with an activated and pro-inflammatory state. By contrast, blood T cells exhibited higher expression of genes associated with quiescence (*TCF7, LEF1*) (Choi et al., 2015) and lymphoid homing (*SELL*) compared to airway T cells (Figure 2E,F). These scRNA-seq results demonstrate compartmentalization of activated TRM populations in the airway of severe COVID-19 in the context of relatively quiescent blood T cells, suggesting that the protective T cell response is targeted to the respiratory environment.

### Resident B cell subsets in the airway of COVID-19 patients

The vast majority of B cells profiled in COVID-19 patients were from the blood; however, there was a small but detectable population in the airways (Figure 1C, 1D). Comparing B cell profiles by flow cytometry analysis revealed differential expression of key B cell markers delineating specific B cells subsets in the airways and blood (Figure S5A, S5B). In particular, airway B cells exhibited increased expression of CD69, a marker expressed by human tissue resident B cells (Weisel et al., 2020), and activation markers CD86 and CD95 (Figure S5A, S5B). Phenograph clustering further delineated subsets of activated and tissue-resident B cells present in airways, while blood contained higher frequencies of CXCR5^+^ and naïve B cells (Figure S5C-E). These results indicate compartmentalization of specific B cell subsets in airway, providing further support for spatial segregation of adaptive immunity.

### Airway monocytes/macrophages exhibit activation and inflammatory profiles

We applied similar high-dimensional flow cytometry and scRNA-seq analysis to the monocyte/macrophage populations in paired airway and blood samples from COVID-19 patients. Phenotypic profiling of airway and blood samples defined a major monocyte/macrophage population (see Figure 1), which largely segregated by site (Figure 3A). Expression of markers HLA-DR, CD11c, and CD16 distinguished airway from blood monocyte/macrophages, while those in blood expressed higher levels of CD14 and CD163 (Figure 3A, B). There was no difference in the relative expression of CD64 and CD86 between airway and blood monocyte/macrophages (Figure 3B). Phenograph clustering of monocytes/macrophages identified 20 clusters, which were coalesced into 6 clusters classified by activation (HLA-DR and CD86) and major monocyte subsets: classical, intermediate, and non-classical (Kapellos et al., 2019) (Figure 3C). Non-classical monocytes/macrophages (both non-activated and activated) and activated classical monocytes/macrophages were enriched in the airway (Figure 3D, E). By contrast, classical and intermediate monocytes/macrophages without activation markers were increased in the blood (Figure 3D, E). These data indicated increased activation of monocyte/macrophage lineages in the airway compared to blood.

**Figure 3.**
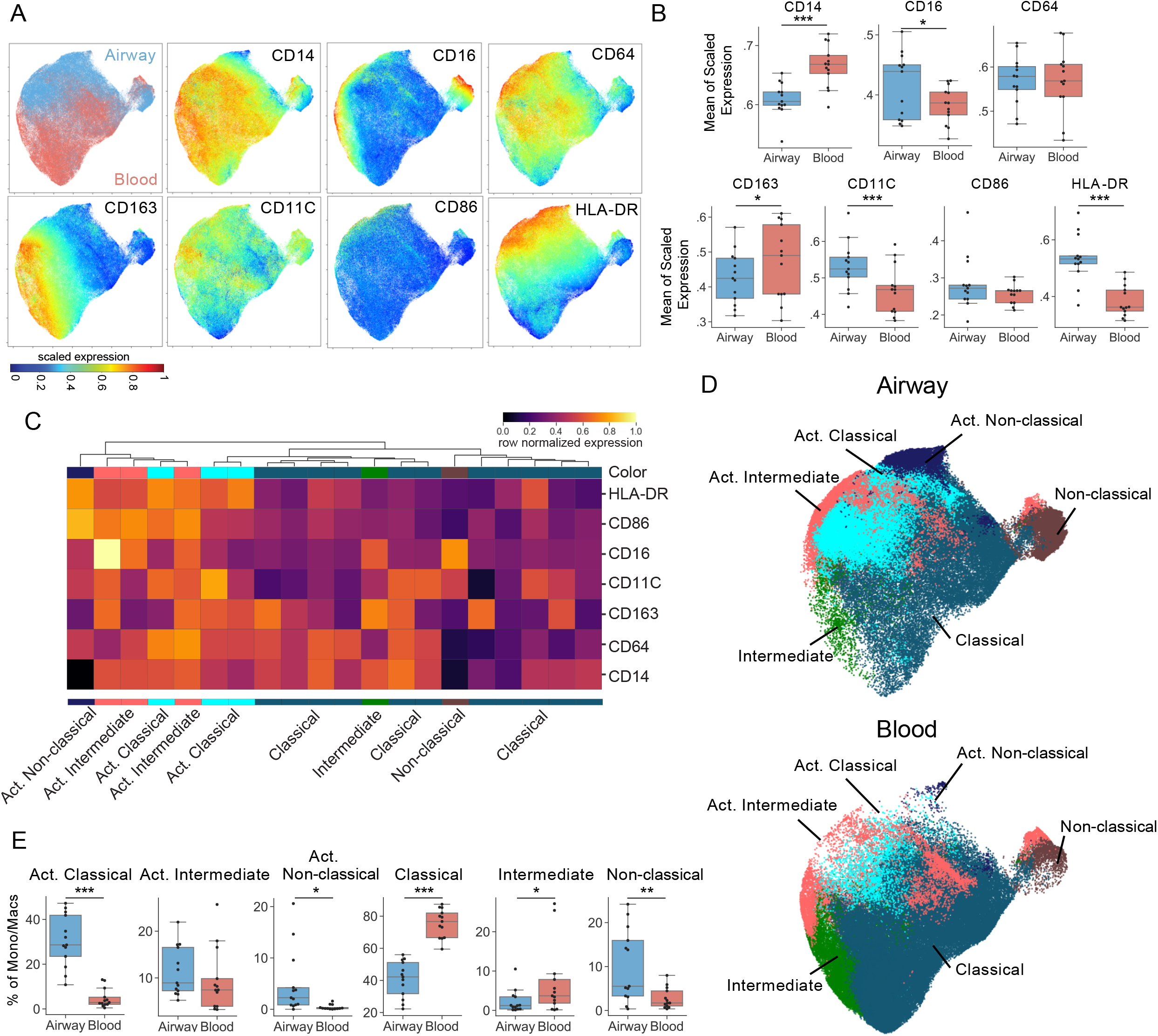
Monocyte/macrophage lineage cells are activated in airway of COVID-19 patients. (A) UMAP embedding of aggregated flow cytometry data obtain in Figure 1 showing expression of major myeloid markers in airway and blood. (B) Mean expression of each myeloid-associated marker within the airway (blue) or the blood (red) samples shown as boxplots with each dot representing individual patient data averaged for all timepoints (C) Heatmap displaying expression of markers within phenograph-generated, hierarchical monocyte/macrophage clusters. The 20 phenograph clusters were collapsed into 6 cellular subsets based on common myeloid nomenclature – classical, intermediate and non-classical, and whether they were activated (“Act.”). Heatmap data are colored by row normalized value for each sample. (D) UMAP embedding of the different subsets colored as in C from airway and blood samples. (E) Boxplots showing compiled frequency of each monocyte/macrophage subset displayed as an average of all time points collected on a per donor basis. Statistical significance was calculated using a paired T-test and indicated by ***, p ≤ 0.001; **, p ≤ 0.01; *, p ≤ 0.05.

We further investigated the subset delineation, differentiation, and functional state of monocyte/macrophages by scRNA-seq. Transcriptionally, airway monocyte/macrophages exhibited certain shared and distinct gene expression patterns compared to blood counterparts, which were consistent across individuals (Figure 4A, Table S6). There was comparable expression of lineage-defining genes, including *CD14, FCGR3A* (CD16), *CD68*, and *CD163*, for monocyte/macrophages in the airway and blood. However, several genes distinguished the two sites, including airway-specific expression of tissue macrophage markers *MARCO* and *MRC2* (CD206) (Bharat et al., 2016) and the integrin *ITGAV* (encoding the vitronectin receptor for tissue matrix interactions), while blood counterparts expressed higher levels of transcripts for chemokine and homing/egress receptors (*CX3CR1, CCR2, SELL, S1PR4*) (Figure 4A). For genes associated with myeloid cell function, airway monocytes/macrophages expressed highly elevated levels of transcripts for pro-inflammatory mediators compared to blood, including chemokines for recruitment of monocyte/macrophages (*CCL2, CCL3, CCL4*), lymphocytes (*CCL18, CCL20, CCL23*), and neutrophils (*CXCL3, CXCL5*), complement components (*C3, C1QB, C1QC*), and matrix metalloproteinases (*MMP9, MMP14*) implicated in tissue damage in ARDS (Hendrix and Kheradmand, 2017) (Figure 4A). Together, these results demonstrate distinct tissue and functional profiles of airway monocyte/macrophages compared to those in the blood. Importantly, airway monocytes/macrophages persist in a highly inflammatory state with elevated expression of chemotactic mediators, suggesting potential roles for lung macrophages in recruiting immune cells to the lung in severe COVID-19.

**Figure 4.**
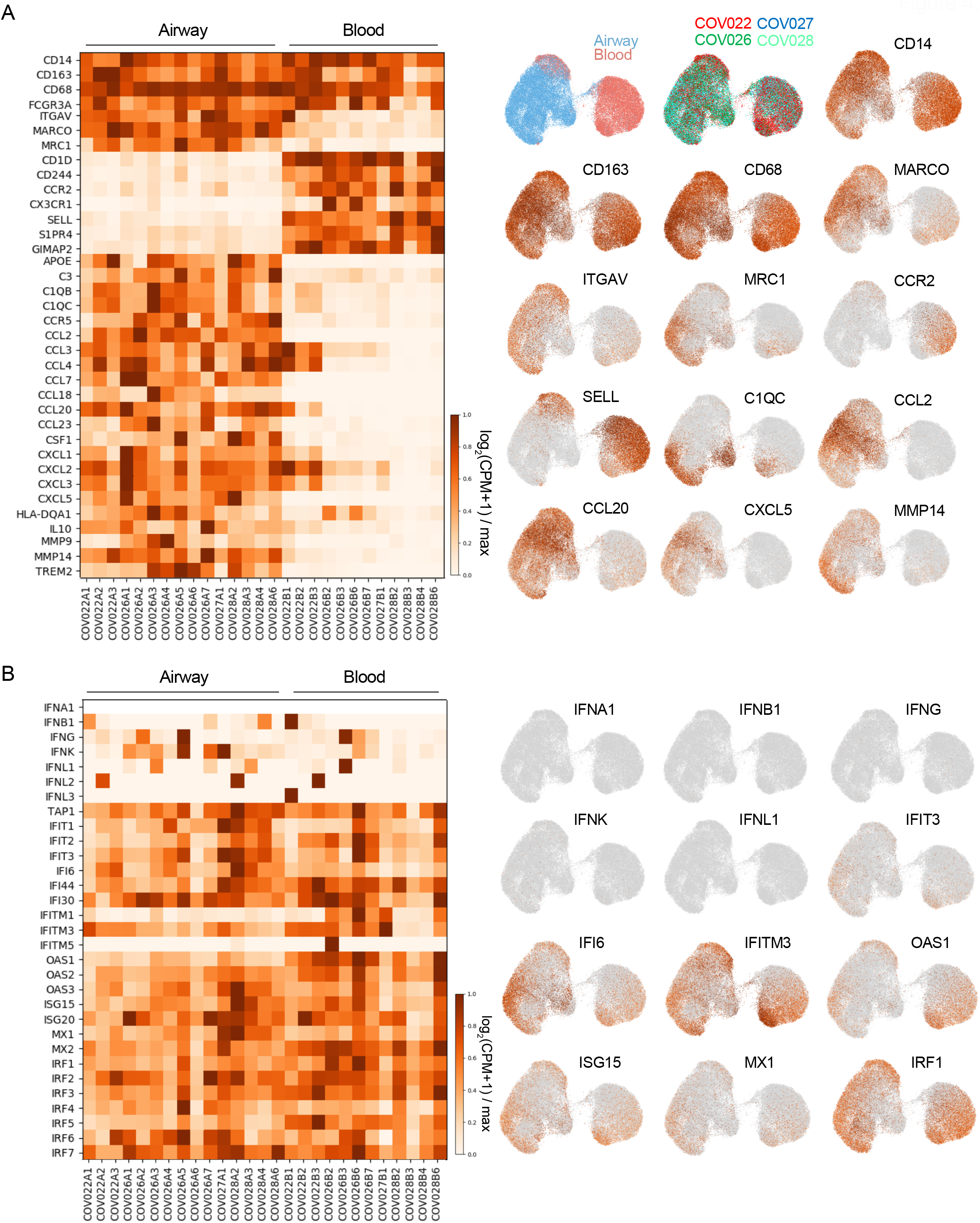
Airway contains tissue macrophages and monocytes with highly inflammatory profiles compared to blood. (A) Monocyte/macrophage profiles in airway and blood were analyzed by scRNAseq. Gene expression analysis of scRNAseq of subset-defining genes, homing receptors and key inflammatory molecules for monocyte/macrophages in airway and blood from each patient sample (left). The heatmap shows genes that are not differentially expressed between airway and blood (*CD14*-*FCGR3A*) and genes are consistently differentially expressed (*ITGAV*-*TREM2*). UMAP embedding of total monocyte/macrophage cells obtained from airway (blue dots) and blood (red dots) compiled from four patient samples (right). (B) Expression levels of the IFN response genes between airway and blood (left). UMAP embedding displaying the expression levels of the IFN response genes in individual cells (right) displayed as log_2_(CPM+1).

Interferon (IFN)-regulated genes are associated with innate anti-viral immunity and may be dysregulated in COVID-19 (Hadjadj et al., 2020). Accordingly, we found negligible expression of genes encoding Type I, Type II, and Type III IFNs from monocyte/macrophages (Figure 4B) or epithelial cells (Figure S6), consistent with the lack of SARS-CoV-2 viral sequences (see methods) in scRNA-seq data from 4 patients. However, transcripts associated with multiple interferon-regulated gene families (i.e., ISG, IFI, IFIT, IRF, MX and OAS) were detected in both the airway and blood monocyte/macrophages (Figure 4B), as well as airway epithelial and T cells (Figure S6), suggesting a persisting anti-viral state in these cells. This expression of IFN-regulated genes may be propagated by *IFNG* expressed by airway T cells (Figure 2, S6). Together these results indicate that the principal innate immune function of myeloid-derived cells in severe COVID-19 is production of pro-inflammatory mediators by airway monocyte/macrophages.

### Compartmentalized production of cytokines and chemokines in airway and blood

We further assessed inflammation in both sites by direct examination of cytokine and chemokine content in airway supernatants and plasma samples from an early (day 1) and later (days 3-7) timepoint for each patient (Table S2). We used a microfluidic chip multiplexed secretome proteomic platform for assessment of soluble mediators from each site with high sensitivity from small volumes (see methods)(Farhadian et al., 2020). Overall, we found major, significant differences in the cytokine and chemokine protein content in the airway compared to plasma, but no significant differences between the two timepoints within a site (Figure 5A, Figure S7A,B). Analytes significantly elevated in airways compared to blood include monocyte/macrophage chemoattractants MCP-1 (CCL2), MIP-1α (CCL3), and MIP1β (CCL4) in all samples, as well as granzyme B, IL-7, and TNF-β associated with T cells and homeostasis (Figure 5A, 5B, Figure S7B). By contrast, in the blood, MCP-1 (CCL2), MIP-1α (CCL3), granzyme B, TNF-β, and IL-7 were undetectable, while MIP-1β (CCL4) was present at variable levels across patients (Figure 5A, B). Both blood and airways contained low and/or variable levels of molecules associated with T cell effector function (perforin, IFN-γ, IL-17, and IL-2), additional innate cytokines (IL-6 and IL-8), and TGF-β, while none of the analytes measured were uniquely expressed by blood and not found in airways (Figure 5A, 5B, Figure S7B). Together, these results show compartmentalized production of pro-inflammatory chemokines and cytokines in the airway with a subset of these detected in blood, suggesting that systemic cytokines may derive from inflammatory processes originating at the infection site.

**Figure 5.**
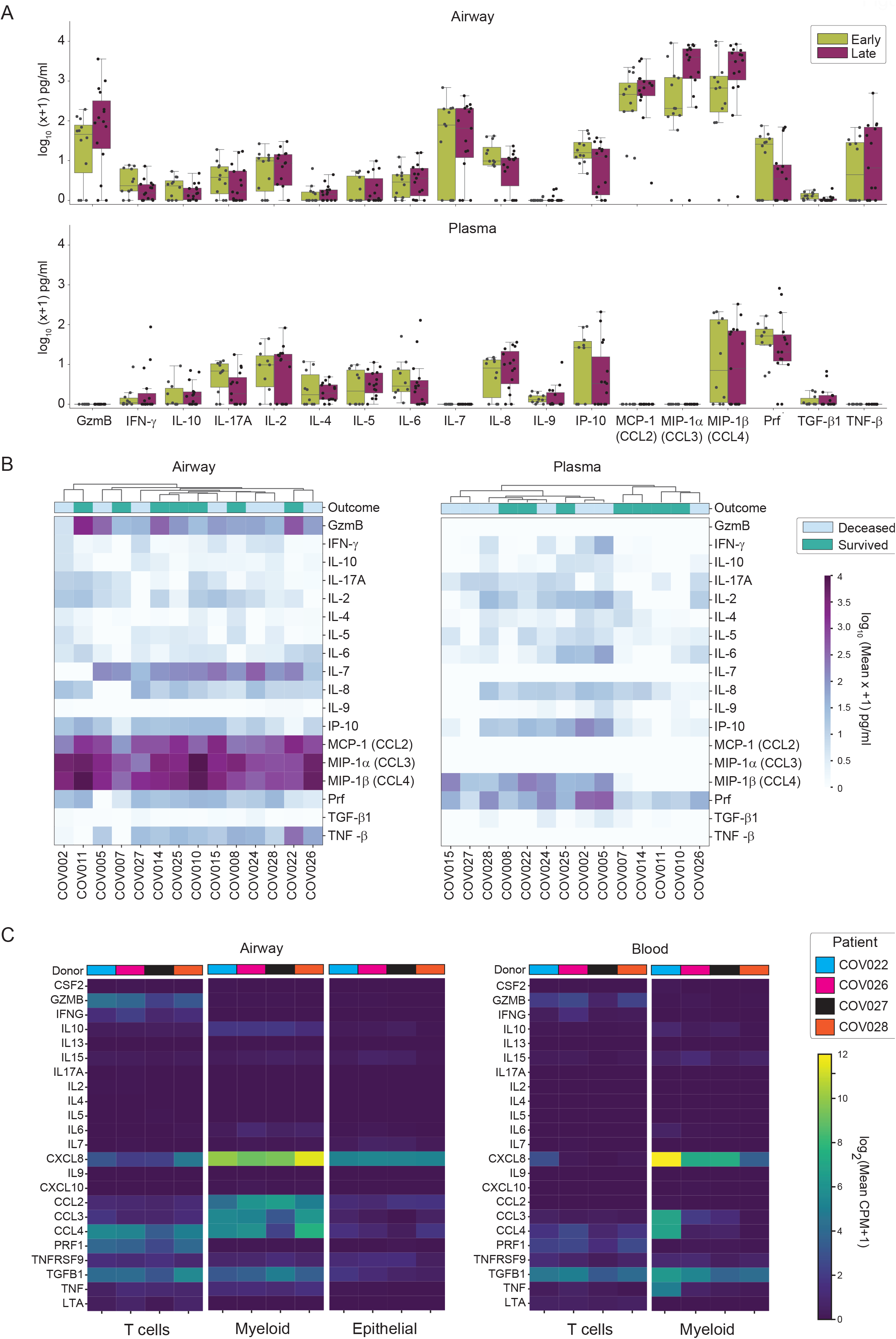
COVID-19 airways contained highly elevated levels of myeloid and T cell-derived cytokines compared to blood. (A) Levels of indicated cytokines and chemokines in the airway (top) and plasma (bottom) compiled from 15 patients depicted in box plots showing log_10_ normalized cytokine expression profiles for an early and late time point (see methods). Each dot represents an individual data point. (B) Heatmap showing log_10_(X+1) pg/mL cytokine levels averaged across both time points in airway (left) and blood plasma (right) samples for each donor. (C) Transcript levels for cytokine expression by major cell lineages identified in by scRNA-seq for each patient samples indicated by color. Heatmap shows log_2_(mean CPM+1) gene expression.

To define the cellular origin of the chemokines and cytokines detected in each compartment, we analyzed transcript expression for each of the analytes from Figure 5B by scRNA-seq. Overall, transcript expression of prominent cytokines/chemokines largely correlated to the protein data; airway myeloid cells expressed high levels of *CCL2, CCL3* and *CCL4* transcripts corresponding to the high levels of the respective proteins in airways, while blood myeloid cells expressed lower or undetectable levels of these transcripts (Figure 5C). Airway and blood myeloid cells also expressed *CXCL8* and *TGFB1*, consistent with the protein data (Figure 5C). In the airways, T cells expressed *GZMB, CXCL8, CCL4, PRF, IFNG*, and *TGFB1* transcripts, which were expressed by blood T cells at lower or variable levels (Figure 5C). Airway epithelial cells expressed predominantly *CXCL8* transcripts, as well as lower levels of transcripts for IL-7 and several chemokines (Figure 5C). Overall, these results demonstrate compartmentalized secretion of monocyte/macrophage-derived chemokines and inflammatory mediators in the airways with potential roles for recruiting immune cells to the lung that may contribute to lung inflammation and tissue damage.

### COVID-19-induced features of airway and blood immune cells

To assess COVID-19-related alterations in airway and blood immune cells that could potentially contribute to disease pathogenesis, we obtained baseline controls of blood from uninfected, healthy adults, and airway washes from lungs of SARS-CoV-2-negative organ donors as done previously (Snyder et al., 2019). High-dimensional flow cytometry analysis of control healthy blood (HB) and airway (HA) samples (Table S7), in conjunction with the COVID-19 patient samples from Figure 1 (COVID-19 blood (CB); COVID-19 airway (CA)) revealed non-overlapping features of COVID-19 and healthy samples for all lineages and particularly within T cells and monocyte/macrophages (Figure 6A, left). Overall immune cell composition and T:monocyte/macrophage cell ratio were similar in healthy and COVID-19 airway samples; however, a dramatic increase in circulating monocyte frequency resulting in a reduced T: monocyte/macrophage ratio was observed in COVID-19 blood relative to healthy controls (Figure 6A, middle and right). By PCA, airway and blood samples were distinct, irrespective of disease; however, healthy and COVID-19 airway samples were intermixed, while healthy blood samples clustered separately from COVID-19 blood samples (Figure 6B). These findings indicate that COVID-19-specific alterations in immune cell composition are manifested more dramatically in blood than in airways.

**Figure 6.**
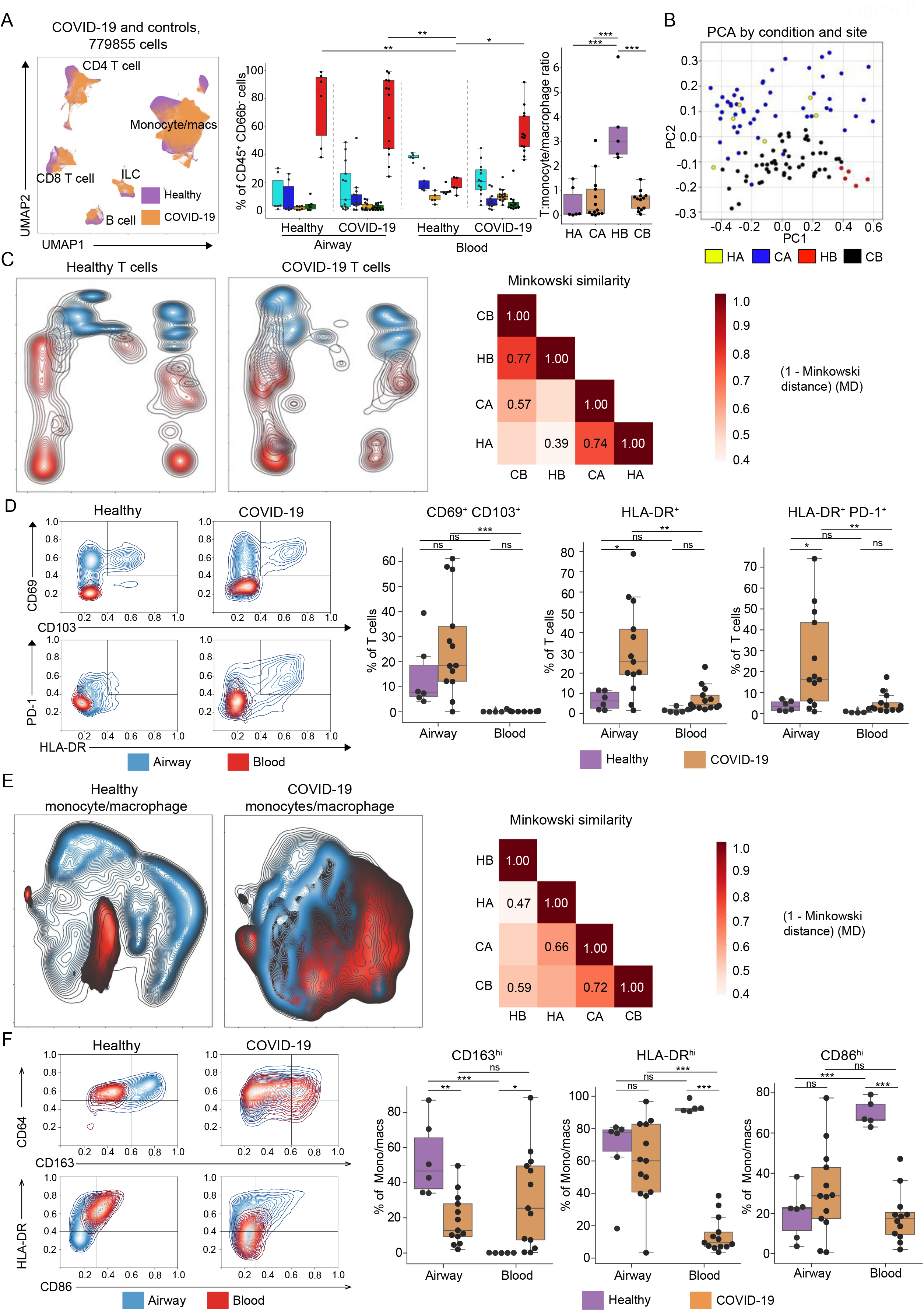
Defining COVID-19-associated immune responses relative to healthy blood and airway samples. Blood was obtained from healthy adults and airway samples from lungs of SARS-CoV-2-negative organ donors; immune cells were stained with the flow cytometry panel in Figure 1 and analyzed in conjunction with COVID-19 patient samples. (A) Comparison of major immune cell lineages in healthy (n=6 airway and 5 blood) and COVID-19 airway (n=54) and blood (n=54) samples. *Left*: UMAP embedding of samples colored by condition (healthy – purple; COVID-19 – orange). *Middle*: Box plots showing the frequency of indicated immune cells from total CD45^+^CD66B^-^ cells for each site. Each dot is the average of all time points per patient/donor. *Right*: Ratio of T: monocyte/macrophage for each site and condition. (B) PCA of mean marker expression (average of each time point for COVID-19 samples) for COVID-19 airway (blue), healthy airway (yellow), COVID-19 blood (black) and healthy blood (red). (C) T cell compartmentalization in airways and blood in health and COVID-19. *Left:* UMAP embedding of the T cell expression data for COVID-19 and healthy controls across airway and blood (upper two panels). *Right*: Correlation heatmaps calculated using Minkowski distance (MD) measures (shown in heat maps as 1-MD) with higher values indicating greater similarity between two samples. HA – healthy airway, HB – healthy blood, CA – COVID-19 airway, CB – COVID-19 blood. (D) Expression of T cell residency and activation markers in airway and blood of healthy and COVID-19 samples. *Left*: Contour plots showing mean expression of indicated markers within the airway (blue contours) and blood (red contours) by condition (healthy or COVID-19). *Right*: Boxplots show frequency of cells expressing indicated markers, for each sample. (E) Monocyte/macrophage compartmentalization in airways and blood in health and COVID-19. *Left:* UMAP embedding of the monocyte/macrophage compartment of COVID-19 and healthy controls across airway and blood (upper two panels). Lower panels indicate correlation heatmap as an average by condition and site (lower right panel). Minkowski distance metric calculated as in D but for myeloid cells. HA – healthy airway, HB – healthy blood, CA – COVID-19 airway, CB – COVID-19 blood. (F) Expression of monocyte/macrophage markers in airway and blood. *Left:* Contour plots showing indicate mean expression of indicated markers by monocyte/macrophages in airway (blue contours) and blood (red contours) samples by condition (healthy or COVID). *Right:* Boxplots indicate percentage of cells within each condition and site that were positive for specific markers, given as an average over all time points for COVID-19 samples. Statistical significance was calculated using a one-way ANOVA followed by a Tukey HSD and indicated by ***, p ≤ 0.001; **, p ≤ 0.01; *, p ≤ 0.05.

To more closely examine site-specific differences between immune cells in healthy and COVID-19 individuals, we analyzed T cell and monocyte/macrophage populations separately. UMAP embeddings of T cells from blood and airway showed compartmentalized profiles for both healthy and COVID-19 samples (Figure 6C, left). We calculated a Minkowski Distance (MD; see methods) to quantify the similarity in T cell populations across conditions and sites, with higher values denoting greater similarity. T cell populations within each site were similar in healthy and COVID-19 samples (HB v. CB MD = 0.77; HA v. CA MD = 0.74), whereas T cell populations in the two sites were more distinct (HA v. HB MD = 0.39, CA v. CB MD = 0.57) (Figure 6C). Comparing T cells in healthy and COVID-19 samples, there were increased frequencies of CD69^+^CD103^+^ TRM and T cells expressing activation markers HLA-DR and PD-1 in the airways of COVID-19 patients compared to uninfected individuals; these markers were not expressed significantly by blood T cells in COVID-19 nor in healthy controls (Figure 6D). The compartmentalized activation of T cells in airways in COVID-19 provides further evidence for dynamic T cell immunity at the infection site.

For monocytes/macrophages, UMAP embeddings revealed compartmentalized profiles between healthy airway and blood, but considerable overlap of monocyte/macrophage profiles between COVID-19 airway and blood (Figure 6E). Accordingly, Minkowski distance calculations confirmed that healthy airway and blood monocyte/macrophage subsets were distinct (MD = 0.47), while in COVID-19 patients airway and blood myeloid cell profiles were more similar (MD = 0.72) (Figure 6E, right). Specifically, CD163, a scavenger receptor typically expressed by tissue macrophages and monocytes in response to inflammation (Buechler et al., 2000), was expressed in control airway macrophages and not by blood monocytes; however, in COVID-19 samples, the proportion of monocytes/macrophages expressing high levels of CD163 (CD163^hi^) was similar in both sites (Figure 6F). Moreover, monocytes in healthy blood samples uniformly expressed HLA-DR and CD86, while monocytes from COVID-19 blood exhibited significantly reduced proportions of HLA-DR^hi^ and CD86^hi^ cells (Figure 6F), consistent with recent findings regarding blood monocyte profiles in severe COVID-19 and suggestive of an immature phenotype (Schulte-Schrepping et al., 2020). Taken together, these results indicate profound alterations in blood monocytes in COVID-19, which also share similar features with airway macrophages, suggesting that airway resident myeloid cells in severe COVID-19 may derive, in part, from these circulating precursors and that interactions between airway and blood myeloid cells may contribute to disease pathology.

### Accumulation of CD163^+^ cells in the lungs of severe COVID-19 patients

We hypothesized that the production of monocyte-chemoattractant chemokines by airway monocyte/macrophages along with the elevated levels of CD163^+^ monocytes in COVID-19 blood may result in their dysregulated infiltration into the lung. We therefore examined immune cells in lung autopsy samples from COVID-19 patients with diffuse alveolar damage, the main pathological finding associated with COVID-19 ARDS (De Michele et al., 2020), relative to lungs from uninfected, deceased organ donor controls (Carpenter et al., 2018) (Table S7). In the airways of uninfected lungs, T cells were clustered around the airway epithelium, while CD163^+^ monocytes/macrophages were dispersed in the parenchyma (Figure 7A, top left). In the lungs of individuals who succumbed to COVID-19 ARDS, there was a marked and dramatic increase in CD163^+^ monocytes/macrophages and damaged airway epithelium that was partially denuded and sloughing off into the lumen (Figure 7A, top right). In particular, CD163^+^ monocytes/macrophages aggregated in the alveolar spaces of COVID-19 infected lungs and not in controls, suggesting their participation in lung damage in COVID-19. Quantitative analysis of the lung imaging data showed significant increases in the frequency and density of CD163^+^ monocytes/macrophages in COVID-19 versus controls, while lymphocyte content was not similarly increased (Figure 7B). We assessed expression of genes associated with cell cycle or proliferation (*Ki67, TOP2A, UBE2C*) in monocyte/macrophage populations in the airway or blood by scRNA-seq, revealing no significant expression of these markers (Figure 7C). Together with the high-dimensional analysis of airway immune cells, these findings implicate the recruitment of immature monocytes from the periphery into the lung, where they subsequently become highly pro-inflammatory and drive the pathogenesis of severe COVID-19.

**Figure 7.**
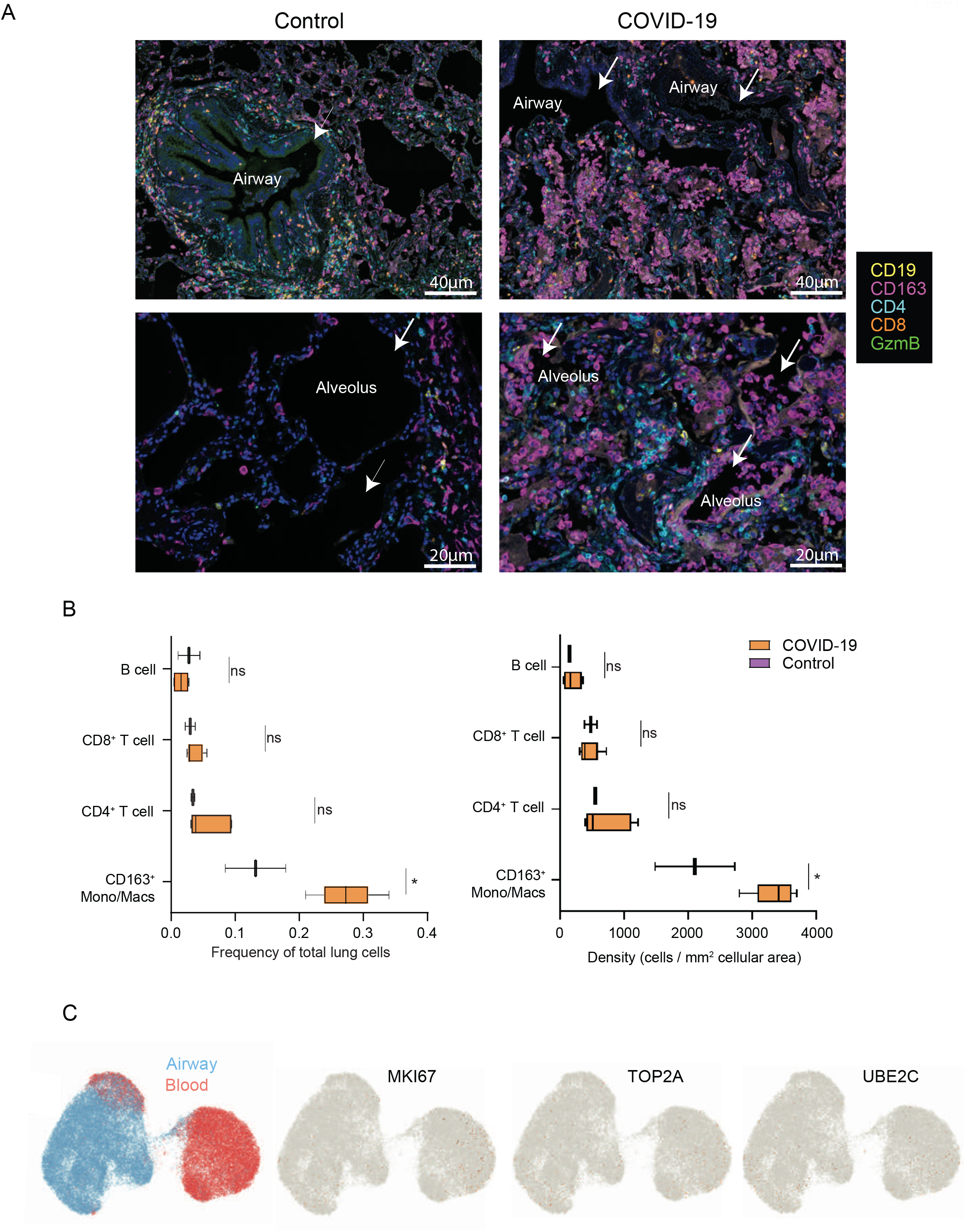
Lung pathology in COVID-19 shows extensive accumulation of CD163^+^ cells associated with cellular recruitment. (A) Lung sections obtained from non-diseased organ donors and autopsy specimens from COVID-19 patients with diffuse alveolar damage were stained with indicated antibodies and analyzed using Vectra. Representative images show T cell (CD4, CD8), and monocyte/macrophages (CD163) staining in the lungs of uninfected controls (left) and COVID-19 patients (right). (B) Quantitation of T cell and monocyte/macrophage content in control (n=2) and COVID-19 (n=5) lungs as a frequency of total lung cells or density (cells per mm^2^ cellular area) using InForm software. Statistical significance indicated by ***, p ≤ 0.001; **, p ≤ 0.01; *, p ≤ 0.05. (C) Expression of genes associated with proliferation by scRNA-seq in monocyte/macrophages derived from airway and blood as in Figure 4.

## DISCUSSION

During the SARS-CoV-2 pandemic, restoration of normal life is impeded first and foremost by the most severe COVID-19 cases, including debilitating ARDS and its high mortality. Numerous studies have now identified characteristic features of innate and adaptive immunity to SARS-CoV-2 infection that are detectable in blood (Kuri-Cervantes et al., 2020; Laing et al., 2020; Lucas et al., 2020; Mathew et al., 2020; Moderbacher et al., 2020; Schulte-Schrepping et al., 2020); however, the initiation, function, and establishment of immune responses for respiratory viruses occur in the lung and respiratory tract. Several studies have separately assessed cellular composition within the respiratory environment in bronchiolar lavage samples and lung autopsies (Damiani et al., 2020; Liao et al., 2020; Veras et al., 2020). Characterizing respiratory immune responses *in situ* in the context of circulating immune cell populations is needed to dissect mechanisms of disease pathogenesis to combat this pandemic.

In this study, we obtained paired respiratory and blood samples from patients with severe COVID-19 longitudinally during the course of intensive care hospitalization. We employed high dimensional profiling by spectral flow cytometry and scRNA-seq as well as multiplex cytokine quantification and immunofluorescence imaging to characterize airway and systemic immune responses and their interactions, revealing key insights into disease pathogenesis. Importantly, we found that innate and adaptive immune responses in severe COVID-19 predominate in the respiratory tract and are qualitatively and quantitatively distinct from immune dynamics in the blood. The most striking differences in immune cells between airways and blood were identified within both T cell and monocyte/macrophage populations. Moreover, increased frequencies of T cells and decreased macrophage/monocyte frequencies exclusively in the airways correlate with better outcome and younger age, further indicating key roles for these cells at the infection site in disease pathogenesis.

T cells in COVID-19 airways were predominately TRM, most of which exhibited features of activation, including surface phenotypes (HLA-DR^hi^PD-1^hi^CD127^lo^) and upregulated expression of transcripts for effector molecules such as perforin, *GZMB* and *IFNG*. This was not the case in the blood of COVID-19 patients, which lacked TRM and activated T cell profiles. Furthermore, activated TRM were detected specifically in airways of COVID-19 patients and not in airway washes of organ donor controls, indicating a virus-directed response, although insufficient T cell numbers in airways precluded direct measurement of SARS-CoV-2-specific T cells. In mouse influenza infection, the presence of activated lung TRM *in situ* to influenza infection correlates with virus-specific responses (Paik and Farber, 2020; Turner et al., 2014), further supporting that *in situ* activation is a surrogate for anti-viral responses. In human SARS-CoV-2 infection, blood may serve as a consistent and reliable indicator for detection of SARS-CoV-2-specific T cells and the establishment of adaptive immune memory (Grifoni et al., 2020; Moderbacher et al., 2020; Weiskopf et al., 2020). However, our results suggest that measuring global T cell activation markers in the periphery, as done in recent studies (Mathew et al., 2020; Takahashi et al., 2020), may not provide an accurate assessment of the virus-targeted immune response *in situ* during active disease.

Airway monocytes/macrophages in COVID-19 patients differed significantly from their blood cell counterparts, with increased frequencies of activated subsets, increased expression of transcripts associated with tissue macrophages (e.g. *MARCO, MRC1, ITGAV)*, and high-level expression of genes encoding pro-inflammatory mediators such as CCL2, CCL3, CCL4, CXCL8, matrix metalloproteases, and complement components. Consistent with this inflammatory profile, excessive levels of MCP-1/CCL2, MIP-1α/CCL3, and MIP-1β/CCL4 protein were detected in the airways, but not in blood, further supporting a role of airway macrophages in initiating and perpetuating the inflammatory responses in severe COVID-19. This phenotypic and functional profile of COVID-19 airway monocytes/macrophages shares features with human macrophages in ARDS due to non-infectious causes, including the production of CCL2 and CXCL8, as well as induction of MMPs and complement (Aggarwal et al., 2014; Morrell et al., 2019). In ARDS, CCL2-expressing airway macrophages recruit inflammatory monocytes expressing the CCL2 binding receptor CCR2, which contribute to lung damage; airway macrophages can subsequently facilitate repair through TGF-β production (Aggarwal et al., 2014). Our results indicate that COVID-ARDS shares some key features with ARDS resulting from other infectious or non-infectious causes.

Our coordinate analysis of airway and blood myeloid cells and soluble mediators suggest an analogous role for airway macrophages driving lung damage in COVID-19 ARDS through recruitment of circulating monocytes. We show that blood monocytes in severe COVID-19 express increased levels of CCR2 transcripts and aberrant CD163^+^HLA-DR^lo^/CD86^lo^ phenotypes compared to healthy blood monocytes. Reduced HLA-DR expression indicative of immature monocytes has been identified in blood myeloid cells in severe COVID-19 (Schulte-Schrepping et al., 2020; Silvin et al., 2020) and may derive from inflammation-induced mobilization of immature monocytes from the bone marrow, termed emergency myelopoiesis (Schultze et al., 2019; Shi et al., 2011; Venet et al., 2020). While a cytokine storm marked by elevated levels of serum cytokines is implicated in pathogenesis of severe COVID-19 and emergency myelopoiesis (Chau et al., 2020; Copaescu et al., 2020; Lucas et al., 2020; Schulte-Schrepping et al., 2020), our results show that inflammatory cytokines detected in the blood lacked CCL2 and other chemokines, which direct recruitment of multiple immune cell types. Our findings rather suggest that pro-inflammatory cytokines emanating from the respiratory tract recruit circulating inflammatory monocytes to the lungs and perpetuate lung damage. Immunofluorescence imaging of lungs from severe COVID-19 patients shows a striking increase in CD163-expressing monocytes/macrophages within the damaged lung tissue that lack proliferative signatures and therefore likely derive from recruitment. These cells specifically accumulate in the alveolar spaces of the lungs, a key site for blood gas exchange, suggesting their involvement in diffuse alveolar damage commonly seen in COVID-19 pathology (De Michele et al., 2020).

Our results defining airway immune responses in COVID-19 and their relation to the corresponding immune reactants in blood have profound implications for treating and preventing disease. Treatments targeting systemic inflammation, either globally with steroids or specifically with cytokine blockade, have shown variable efficacy in severe COVID-19 (Della-Torre et al., 2020; Furlow, 2020). Our results suggest that targeting airway-derived cytokines such as CCL2 through CCR2 antagonists or other airway-specific mediators may be more effective in reducing lung damage or even promoting recovery from ARDS in severe COVID-19. A similar role for CCL2-mediated monocyte recruitment in lung pathology was demonstrated in mouse models of influenza infection (Lin et al., 2008; Lin et al., 2011), suggesting a generalized mechanism for respiratory virus-induced lung injury. Because our scRNA-seq analysis showed that the elevated airway mediators derive chiefly from the lung macrophages, treatments which regulate these cells may also mitigate the clinical course of disease.

Our finding that increased proportions of airway T cells are associated with better outcome and younger age suggests that promoting lung-localized immune responses is an important consideration for vaccine design. In mouse models, intranasal administration of the live-attenuated influenza vaccine or bacterial-based vaccines can promote establishment of lung TRM that mediate protective immunity to pathogen challenge (Allen et al., 2018; Zens et al., 2016). Intravenous administration of the BCG vaccine to non-human primates was recently shown to generate substantial populations of lung TRM, which correlated strongly with protection from tuberculosis (Darrah et al., 2020). The current SARS-CoV-2 vaccines in phase III trials target generation of neutralizing antibodies and are robust strategies for establishing sterilizing immunity (Jeyanathan et al., 2020); however, respiratory targeting could be considered for individuals who are unable to develop effective antibody responses. These cohorts may include the immunocompromised or the elderly, or this strategy could be used as a booster for those at risk for infection due to frequent interactions with others through their living or work situations. Indeed, a recent pre-clinical study demonstrated that intranasal administration of a recombinant SARS-CoV-2 vaccine promoted lung TRM generation and protection from viral challenge in a mouse model (Hassan et al., 2020).

In summary, our study provides a dynamic view of ongoing respiratory immunity in severe COVID-19, revealing compartmentalization of protective and pathogenic immune responses in the lung. These findings have important implications for how we monitor, treat and protect from this pandemic and future infectious challenges to the respiratory tract.

## Data Availability

The scRNA-seq data for each sample including count matrices, normalized counts, metadata, cell annotations, and UMAP embeddings are available on the COVID-19 Cell Atlas along with interactive visualizations with the url provided below.

https://www.covid19cellatlas.org/index.patient.html

## Acknowledgements

We wish to express our gratitude to the Medical ICU nurse champions, Cora Garcellano, Tenzin Drukdak, Harriet Avila Raymundo, Lori Wagner, and Ricky Lee, who led the efforts to obtain patient samples for the adult ARDS patients, to Evelyn Hernandez and Lorena Gomez for their roles as clinical coordinators, and to the nurses and clinical staff in the Pediatric Intensive Care Unit of MSCHONY. We acknowledge the dedication, commitment, and sacrifice of the other nurses, providers, and personnel who helped care for these patients during the COVID-19 crisis. We acknowledge the suffering and loss of our COVID-19 patients and of their families and our community. We also gratefully acknowledge the generosity of the donor families and the exceptional efforts of LiveOnNY transplant coordinators and staff for the donor lungs.

This work was supported by NIH grants AI128949 and AI06697 awarded to D.L.F., a Chan Zuckerberg Initiative COVID-19 grant to D.L.F. and P.A.Sims, and an R01AI093870 awarded to A.J.Y. P.D. was supported by a CRI-Irvington Postdoctoral Fellowship and P.A.S. by a Canadian Institutes of Health Research Fellowship. T.J.C. is supported by NIH K23 A1141686 and S.P.W. is supported by NIH K08 DK122130. Research reported in this publication was performed in the Human Immune Monitoring Core, the Columbia Single Cell Analysis Core, and the Sulzberger Columbia Genome Center, which are supported by an NCI cancer center support grant P30CA013696. The content is solely the responsibility of the authors and does not necessarily represent the official views of the National Institutes of Health. We thank Eldad Hod for use of his laboratory for sample processing, and Carly Ziegler and Dr. Alex Shalek of MIT for sharing their merged human / SARS-CoV-2 genome and transcriptome annotation.

## Author Contributions

P.A.Sz., S.W., J.G., P.D. processed samples, designed and optimized high-dimensional flow cytometry panels, analyzed data, made figures, and wrote the manuscript. P.A.Sz. and S.W. processed samples for scRNAseq profiling and encapsulation using 10X Chromium; I.K. prepared and sequenced the 10X libraries. P.D. designed the Python pipeline for flow cytometry data. M.B. monitored and consented ICU patients, oversaw clinical data analysis, and collected samples. T.C. obtained and maintained IRB protocols, consented patients, and processed samples. M.M.L.P., R.M., E.I., M.C. obtained and processed patient samples. S.E.M., C.P., and A.JY. statistically analyzed longitudinal data; J.D.-P. captured and analyzed patient data; J.Z., M.S. S.M. performed cytokine analysis of airway supernatants and blood plasma; S.P.W. planned, designed, and analyzed lung autopsy imaging experiments; A.S. provided lung autopsy samples and associated data; A.K. performed immunohistochemistry of lung autopsies. P.A.Sims planned scNAseq experiments, analyzed data, and, wrote and edited manuscript. D.L.F. oversaw compliance, planned experiments, coordinated sample acquisition and data acquisition/analysis, analyzed data, and wrote and edited the paper.

## Declaration of Interests

J.Z., M.S. and S.M. have competing interests with IsoPlexis. The remaining authors declare no competing interests.

## Supplemental Figure Titles and Legends

**Figure S1. Gating strategy for flow cytometry analysis** (A) FACS plots denoting the gating strategy used for analysis of myeloid and lymphocyte populations using the Aurora flow cytometry from complex populations in airway samples, with complementary gating for blood cells. Total cells were initially gated on CD45^+^ cells versus time to eliminate non-hematopoietic cells and debris; doublets were excluded followed by exclusion of neutrophils (FSC-A^hi^ CD66b^+^). Finally, dead cells were excluded by gating on CD45 and LIVE/DEAD blue. The resulting populations contained the full complement of mononuclear immune cells used for downstream analysis.

**Figure S2. Expression of lineage defining markers determined by flow cytometry and scRNA-seq** (A) UMAP embedding displaying expression of lineage-defining markers for the major immune cell subsets in combined airway and blood samples from 13 COVID-19 patients. Heatmap of scRNA-seq data displaying gene expression of lineage defining markers for both the airway and blood across the four donors. Heatmap data are colored by log_2_(CPM+1)/max values for each sample.

**Figure S3. Major immune cell lineages over time in COVID-19 patients**. (A) Individual patient data displaying the proportion of each major immune cell lineage over the time course of sample collection. D – deceased, S – survived. (B) Classification performance of longitudinal K-means clustering for different combinations of immune cell trajectories. The percentage of donor outcomes that were successfully classified as deceased or survived is shown when all subsets, only myeloid and T cells, only myeloid, or only T cells were used for clustering. Colors denote whether airway, blood, or both airway and blood trajectories were included. (C) Association of ILC, B cells, T:myeloid and CD4:CD8 content in airway (left) and blood (right) with outcome (deceased or survived) and correlation with age. Statistical significance was calculated using Mann-Whitney U-tests (box-plots) or Pearson correlations (scatter plots) and indicated by ***, p ≤ 0.001; **, p ≤ 0.01; *, p ≤ 0.05.

**Figure S4. T cell marker expression in the airways and blood of COVID-19 patients**. (A) UMAP embeddings indicate site of origin for total T cells in the airway and blood of COVID-19 patients (top left) and expression of indicated T cell markers. (B) Boxplots showing mean of scaled expression of T cell markers on total T cells for each patient averaged across all timepoints, Statistical significance was calculated using a paired T-test and indicated by ***, p ≤ 0.001; **, p ≤ 0.01; *, p ≤ 0.05.

**Figure S5. B cell subsets in airway and blood** (A) UMAP embedding of the expression of key B cell markers in airway and blood samples. (B) Boxplots showing mean of scaled expression of B cell markers on total B cells for each patient averaged across all timepoints, (C) Heatmap displaying expression of markers within phenograph-generated clusters for B cell subsets. The 12 phenograph clusters were collapsed into 9 subsets designated on the bottom row. Heatmaps are colored by row normalized expression values. (D) UMAP embedding of 9 B cell subsets in the airway and blood. (E) Boxplots showing frequency of each B cell subset among total B cells for each patient averaged across all timepoints, Statistical significance was calculated using a paired T-test and indicated by ***, p ≤ 0.001; **, p ≤ 0.01; *, p ≤ 0.05.

**Figure S6 related to Figure 4. IFN and IFN-related gene signature in COVID-19 airways**. Heatmap showing log_2_(CPM+1) expression of *IFN* and *IFN*-related genes by the indicated airway cell types as determined by scRNA-seq for each patient sample, indicated by color.

**Figure S7. Airway is the major site for production of inflammatory cytokines and chemokines in COVID-19 patients**. (A) Box plots showing log_10_(X+1) normalized cytokine expression profiles in the airway wash (left) and blood plasma (right) samples for an early and late time point collected from 15 donors. Each dot represents an individual data point. (B) Pairwise comparison of cytokine levels averaged across both timepoints in airway wash and blood plasma samples collected from 15 donors. A p-value of < 0.05 was considered significant. For figures, p-value < 0.05 = *, p-value < 0.01 = ** and p-value < 0.001 = **.

## Methods

## LEAD CONTACT AND MATERIALS AVAILABILITY

Further information and requests for reagents should be directed to and will be fulfilled by lead author Donna L. Farber

## Materials Availability Statement

This study did not generate new unique reagents.

## EXPERIMENTAL MODEL AND SUBJECT DETAILS

### Human samples

We recruited patients from and CUIMC/NYP and Morgan Stanley Children’s Hospital of NY with severe COVID-19 and ARDS (n=15) who tested positive for SARS-CoV-2 by polymerase chain reaction (PCR) from nasopharyngeal swabs (Table S1, S2). Blood and airway sampling began within 24-36 hours for all patients. ARDS was defined by clinical consensus criteria; including infiltrates on chest radiograph and a PaO2/FiO2 ratio of less than 300, or pediatric criteria equivalent (Khemani et al., 2015; Ranieri et al., 2012). Sequential Organ Failure Assessment (SOFA) scores were calculated on all hospitalized patients using previously validated adult and pediatric score tools to provide additional clinical insight into patient disease severity (Matics and Sanchez-Pinto, 2017; Singer et al., 2016; Vasilevskis et al., 2016). All patients and samples in this study were enrolled on protocols approved by the Institutional Review Board at CUIMC. Due to the limitations placed on direct contact with infected patients and a need to conserve personal protective equipment, verbal informed consent was obtained from surrogates of critically ill COVID-19-ARDS patients. Healthy blood was obtained from 5 adult volunteers 31-57 years.

Control, non-diseased lung tissues were obtained from deceased organ donors as part of organ acquisition for clinical transplantation through an approved protocol and material transfer agreement with LiveOnNY as described previously (Carpenter et al., 2018; Dogra et al., 2020). Donors were free of cancer, chronic diseases, seronegative for hepatitis B, C, and HIV, and negative for SARS-CoV-2 by PCR (Table S7). Use of organ donor tissues does not qualify as “human subjects” research, as confirmed by the Columbia University IRB as tissue samples were obtained from brain-dead (deceased) individuals.

### Processing of blood samples and isolation of PBMCs from COVID-19 patients

Whole blood collected in heparinized vacutainers was centrifuged at 400 x g for 10 min at room temperature (RT) to isolate plasma, which was then stored at −80 °C for subsequent analysis. PBMCs were isolated using Ficoll-Paque PLUS (GE) density gradient centrifugation in a Biosafety Level 2+ facility. To remove neutrophils, blood was incubated with RosetteSep Granulocyte Depletion Cocktail (Stemcell Technologies), diluted 1:3 in room temperature DPBS, layered over Ficoll-Paque PLUS in 50mL conical tubes, and centrifuged for 20 min at 1,200 x g. The PBMC layer was isolated according to the manufacturer’s instructions. Cells were washed twice with DPBS before counting with the automated NucleoCounter NC-3000 cell counter (ChemoMetec).

### Processing of airway samples and isolation of airway MNCs from COVID-19 patients

To collect airway supernatants, DPBS was added 1:1 directly to airway samples and centrifuged at 400 x g for 10 min at RT. The resulting supernatants were stored at −80 °C for subsequent analysis. To isolate airway MNCs, samples were treated with Benzonase (Millipore Sigma), purified through 100 µm filters, and centrifuged on a density gradient using Ficoll-Paque PLUS. The MNC layer was isolated according to the manufacturer’s instructions. Cells were washed twice with DPBS before counting with the automated NucleoCounter NC-3000 cell counter (ChemoMetec).

### Cell preparation for scRNA-seq, library generation and sequencing

Airway and blood MNC populations were isolated as above, and the remaining neutrophils and red blood cells were removed by incubating samples with biotinylated anti-CD66b and anti-CD235ab antibodies, and depleting antibody-bound cells with streptavidin-coated magnetic beads (Bangs Labs). Dead cells were subsequently removed using the Dead Cell Removal kit (Miltenyi Biotec). The Next GEM Chromium Controller (10x Genomics) and Chromium Next GEM Single Cell 3’ Reagent kit v3.1 (10x Genomics) was used for co-encapsulation and scRNA-seq library construction as per manufacturer’s suggested protocols. Libraries were sequenced on an Illumina NovaSeq 6000, targeting ∼300M raw reads per sample (∼60,000 raw reads per cell). Sample details and number of cells sequenced in each are shown in Table S3.

### Isolation of airway washes from non-diseased lungs

Non-diseased lungs were obtained from deceased organ donors as described above. Airway washes were obtained by flushing out the major airway with 60 mL saline as described (Snyder et al., 2019). Cells were pelleted by centrifugation, resuspended in D-PBS and stained with antibodies for flow cytometry.

### High Dimensional Flow cytometry

For high parameter analysis using the Cytek Aurora panel, 5×10^6^ cells from each site were stained in 5 mL U-bottom tubes in the dark using the following antibody panel; Anti-Human HLA-DR-BUV395, Anti-Human CD16-BUV496, Anti-Human CD163-BUV563, Anti-Human CD33-BUV615, Anti-Human PD-1-BUV661, Anti-Human CD56-BUV737, Anti-Human CD64-BUV805, Anti-Human CCR7-BV421, Anti-Human CD86-SB436, Anti-Human CD28-eFluor 450, Anti-Human CD8-BV480, Anti-Human CD20-Pacific Orange, Anti-Human CD3-BV510, Anti-Human CD45RA-BV570, Anti-Human CD25-BV605, Anti-Human CD27-BV650, Anti-Human CD69-BV711, Anti-Human CXCR5-BV750, Anti-Human CD335-BV785, Anti-Human CD103-BB515, Anti-Human CD66b-FITC, Anti-Human CD14-Spark Blue 550, Anti-Human CD45-PerCP, Anti-Human CD57-PerCP-Cy5.5, Anti-Human TCR gamma/delta-PerCP-eFluor 710, Anti-Human CD1338-PE, Anti-Human CD4-eFlour 568, Anti-Human CD123-PE-CF594, Anti-Human CD95-PE-Cy5, Anti-Human CD11c-PE-Cy7, Anti-Human CD19-Spark NIR, Anti-Human CD127-APC-R700, Anti-Human KLRG1-APC/Fire 750, Anti-Human FoxP3-Alexa Fluor 647. Briefly, cells were washed with DPBS, re-suspended in 1 mL of viability dye and incubated at RT in the dark for 10 min. Following incubation, cells were washed once with cold FACS-buffer (DPBS + 2% FBS + 0.1 mM EDTA) and re-suspended in 200 µl FASC-buffer + 10 µl human TrueStain FcX + 10 ul of True-Stain Monocyte Blocker and incubated in dark for 15 minutes. Following incubation, cells were washed once with cold FACS-buffer and stained in a two-step process. First, the cells were resuspended in a cell-surface marker staining cocktail and incubated on ice for 20 min. For intracellular staining, surface stained cells were fixed for 25min at RT in fixing buffer (Invitrogen cat# 00-5123-43), followed by staining in permeabilization buffer (Invitrogen cat# 00-8333-56) at RT for 30 min. Cells were washed and data was collected on 5-lazer Cytek^®^ Aurora machine (Cytek Bio).

### Highly-multiplexed CodePlex chip secretome proteomics

Cryopreserved tracheal washes and plasma were thawed at room temperature for 30-60 minutes and mixed well by pipetting up and down prior to loading. An aliquot of 5.5 µL of each sample was pipetted into each macrochambers of a CodePlex chip pre-patterned with a complete copy of a 23-plex antibody array. 2% BSA/PBS was used as background control. The chip was then loaded into an IsoLight automation system and various proteins were measured by fluorescence ELISA and analyzed by the IsoSpeak software using the IsoPlexis Human Adaptive Immune Panel: GM-CSF, Granzyme B, IFN-γ, IL-10, IL-13, IL-15, IL-17A, IL-2, IL-4, IL-5, IL-6, IL-7, IL-8, IL-9, IP-10, MCP-1, MIP-1α, MIP-1β, Perforin, sCD137, TGF-β1, TNF-α, TNF-β.

### Multispectral staining and imaging of lung tissue

Representative samples of lung tissue 0.5–1.0 cm in thickness were recovered from organ donors and autopsy cases of individuals diagnosed with COVID-19 and found on post-mortem exam to have pathological findings consistent with diffuse alveolar damage (Table S7). Samples were fixed in 10% formalin (Anatech Ltd.) for 48 hours prior to dehydration and embedding in paraffin. These Lung samples were sectioned at 5-mm thickness and stained using 7-color multispectral Opal reagents (Anti-human CD19-Opal 540, Anti-human CD8-Opal 690, Anti-human CD163-Opal 650, Anti-human CD4-Opal 520, Anti-human GzmB-Opal 570, Anti-human CD3-Opal 620) (Akoya Biosciences, Cat# NEL811001KT) as previously described (Gartrell et al., 2018; Weisberg et al., 2019). The multiplex panel included DAPI (BioLegend cat# 422801) for nuclear counterstaining, CD4 (1:150 dilution), CD8 (1:600 dilution), CD163 (1:200 dilution), granzyme B (GzmB) (1:200 dilution), CD19 (1:50 dilution), CD3 (1:500 dilution). Single controls and an unstained slide were stained with each group of slides. After staining, the sections were mounted in Vectashield Hard Set mounting media (Vector Labs, Cat#H1600) and stored at 4^0^C for up to 48 hours prior to image acquisition. Multispectral imaging and acquisition at 20x magnification (numerical aperture 0.75) was performed using the integrated Vectra 3automated quantitative pathology imaging system (PerkinElmer) as previously described(Weisberg et al., 2019). Images were analyzed using inForm software (PerkinElmer). Representative areas (10-30) from each donor were chosen for quantitative analysis.

## Data Analysis

### Flow cytometry analysis

Flow cytometry data was pre-gated to exclude any doublets, dead cells and CD66b^+^ granulocytes using FlowJo v 10.7 (Tree Star) (Figure S1). Cleaned data was exported as .fcs files with compensated parameters and analyzed further and visualized using a Python (v3.7) (Python Software Foundation. Python Language Reference, version 2.7.) computational pipeline. In brief, first the data was filtered to remove any noise using quantile gates; events that fell below 0.01% of marker expression intensity were removed from the sample. Following initial filtering, data from COVID-19 and healthy samples was merged after subsetting 70,000 events from each sample. Any sample with fewer than 1000 events was removed from further analysis. The merged dataset as was transformed using arcsinh function from Python *numpy* library(van der Walt et al., 2011) after manually adjusting the cofactor for each marker. Following normalization, the dataset was normalized on a 0-1 feature scale for each marker using MinMaxScaler function from Python *scikit-learn* library (Pedregosa et al., 2011). The cleaned, transformed and scaled dataset was used to run the first round of *Uniform Manifold Approximation and Projection* (UMAP) (McInnes et al., 2018) dimensionality reduction to remove any residual granulocyte contamination identified as clusters of CD45^lo^CD66b^+^ cells. The resulting “no neutrophil dataset” dataset was split into COVID-19 and healthy samples and used for downstream analysis.

For further analysis we downsampled the “no neutrophil COVID-19 dataset” to include 20,000 events from each of 141 longitudinal samples and was used to run PCA analysis at sample level using mean expression of markers in each sample, PCA loadings provided in Table S3. We ran UMAP dimensionality reduction (k = 60) on this dataset using 14 lineage-defining markers (CD11c, CD14, CD16, CD19, CD27, CD3, CD4, CD8, CD64, CD56, CD33, CD335, CXCR5, HLA-DR). The data were projected in 2-dimensions using UMAP embeddings and clusters of major immune cell types (CD4 and CD8 T cells, B cells, NK/ILC and Monocytes/Macrophages) were identified based on expression of lineage defining markers (Figure S2B). The frequency of each lineage was averaged for individual donor-site group across all time points and used for hierarchical clustering of samples using “ward” method and “jensenshannon” metric.

For lineage specific analysis, we ran UMAP dimensionality reduction and subsequent *Phenograph* clustering (Levine et al., 2015) on each lineage specific dataset using cell subset defining markers selected based on literature review. Markers used for T cell UMAP are shown in Figure S4A. B cell markers used are shown in Figure S5A and monocyte/macrophages shown in Figure 3A. Major cell subset clusters were identified and functionally similar subsets were coalesced and manually annotated. Heatmaps were generated for average marker expression in each cluster. Data are presented as row normalized expression of marker across all clusters.

For analysis of COVID-19 and healthy samples, paired blood and airway samples across all timepoints from COVID-19 donors were downsampled to 5,000 events, and each healthy airway and blood sample was downsampled to 30,000 events and merged to create a reduced “no neutrophil Healthy + COVID-19 dataset”. UMAP dimensionality reduction and identification of major cell lineages was done as described above for the COVID-19 dataset. To evaluate similarity of samples by condition-site i.e. healthy-blood, healthy-airway, COVID-19-blood and COVID-19-airway we calculated Minkowski distance metric (MD) (Li et al., 2011) for the samples on scaled marker expression values for individual lineages using Python scipy library (Jones et al., 2001). Data are presented as 1-MD (Minkowski similarity) on the heatmap; higher numbers indicate increased similarity and lower numbers indicate reduced similarity between samples. All graphs were generated using the Python *matplotlib* and *seaborn* libraries (Hunter, 2007).

### Classifying donor outcomes using longitudinal K-means clustering

Donors were partitioned into two groups using a longitudinal K-means algorithm applied to the trajectories of the frequencies of myeloid, B cell, CD4 and CD8 T cell and ILC frequencies in blood and/or airways. The proximity of two donors’ trajectories was defined using the sum of the squared Euclidean distances between their subset frequencies at each location at each timepoint, after normalizing each subset frequency across all donors and timepoints. The clustering outcome was robust to this definition of distance, giving identical results when performed using log- or logit-transformed frequencies. Classification performance was defined as the percentage of donors that were assigned to the correct outcome cluster (i.e. deceased or survived). We compared the abilities of different immune cell subsets to distinguish donor outcome by repeating the clustering analysis on different combinations of trajectories. Greater classification performance indicated increased power to identify donor outcomes. Clustering analyses were conducted in *R* version 3.5.3 using the *kml3d* package, version 2.4.2, and the results were visualized using the ggplot2 package.

### Processing of scRNA-seq Data

We used kallisto v0.46.2 in “BUS” mode to pseudo-align the raw reads for each sample to a merged human GRCh38 (Ensembl 93)/SARS-CoV-2 transcriptome (Bray et al., 2016; Kim et al., 2020; Melsted et al., 2019a; Melsted et al., 2019b). To correct for index swapping, which can occur on the Illumina NovaSeq 6000, we applied the algorithm of Griffiths *et al* (Griffiths et al., 2018) to the equivalence classes obtained from kallisto pseudo-alignment. We generated a raw count matrix from the swap-corrected BUS file using bustools v0.40.0(Melsted et al., 2019b), filtered using the EmptyDrops algorithm (Lun et al., 2019), and removed all cells with mitochondrial pseudo-alignment rates >20% or counts per gene greater than two standard deviations above the mean for each sample.

### scRNA-seq Cell Annotation

We merged the scRNA-seq data from all of the airway samples and identified likely markers of specific subpopulations using the previously described drop-out score method for finding genes that are detected in fewer cells than expected given their expression level (Levitin et al., 2019; Szabo et al., 2019). Next, we computed a cell-by-cell Spearman’s rank correlation matrix using these putative marker genes. Using this matrix, we constructed a k-nearest neighbor’s graph (k=20) as input for Louvain community detection as implemented in Phenograph (Levine et al., 2015). To associate the resulting clusters with major cell populations in the airway, we examined the statistical enrichment of the following marker genes in each cluster using the binomial test as described in Shekhar *et al* (Shekhar et al., 2016): T cells (*CD3D, TRAC, TRBC1, TRBC2, TRDC, TRGC1, TRGC2*), NK cells (*NCAM1*), myeloid cells (*CD14, FCGR3A, CD163*), epithelial/club/goblet cells (*EPCAM, SCGB1A1, MUC5B, KRT78*), ionocytes (*CFTR*), neutrophils (*CD16B*), plasma cells (*CD19, JCHAIN*), B cells (*CD19, MS4A1*), platelets (*ITGA2B, PF4*), mast cells (*KIT*), dendritic cells (*FCER1A, CD1C*) and red blood cells (*HBA1, HBA2, HBB*). We identified clusters as likely multiplets based on co-expression of multiple marker sets (e.g. clusters enriched in both *CD14* and *CD3D* were marked as likely T cell / myeloid cell multiplets). All of the cells in these clusters were marked as multiplets.

In the main text, we present focused analyses on myeloid cells, T cells, and epithelial/club/goblet cells from the airway. To further refine our annotation, we re-clustered the cells annotated as each of these three cell types separately using the methods described above. We then re-analyzed the enrichment of cell type-specific markers in the resulting new clusters. As expected, this focused re-analysis of each of these three major populations identified additional putative multiplet clusters and cells that we likely mis-clustered in the initial merged analysis. We conducted two rounds of re-clustering for each of these three major cell types to produce a refined annotation. The top of Figure S2B shows a gene expression heatmap for key markers genes in the merged airway data set colored by patient and cell type annotation. We repeated the above procedure for the merged blood scRNA-seq profiles including a focused re-clustering analysis of the cells that we originally annotated as myeloid and T cells for refinement. The bottom of Figure S2B shows a gene expression heatmap for key markers genes in the merged blood data set colored by patient and cell type annotation.

### scRNA-seq Visualization and Differential Expression Analysis

We generated merged UMAP embeddings for the blood and airway T cells (Figure 2E, F) and the blood and airway myeloid cells (Figure 4). In each case, we first identified genes that were likely to contaminate either the myeloid or T cell profiles in either the blood or airway to avoid including them in any of our downstream clustering, visualization, or differential expression analysis. We conducted pairwise differential expression analysis between all of the cells annotated as a cell type-of-interest (e.g. myeloid) and each group of cells with a different annotation for the blood and airway from each patient separately. For each pairwise comparison, we randomly subsampled the two groups of cells to the same cell number. Next, we randomly subsampled the molecular counts for cells in the two groups such that they have the same average number of counts per cell. We then generated a merged count matrix for the two groups and applied the pooled normalization technique from the *scran* package of Lun *et al* using the *computeSumFactors* function (Lun et al., 2016). Finally, we conducted a gene-by-gene, non-parametric differential expression analysis using the Mann-Whitney U-test as implemented with the function *mannwhitneyu* from the Python package *scipy*. We corrected the resulting p-values for false discovery using the Benjamini-Hochberg Procedure with the function *multipletests* from the Python package *statsmodels*. Using the results of pairwise differential expression analysis, we generated a blacklist of genes for a given cell type by taking any gene with at least 10-fold enrichment in a different cell type with FDR<0.001 in at least two patients. We removed all genes with any enrichment in the cell type-of-interest with FDR<0.001 in any patient to avoid eliminating patient-specific markers of the cell type-of-interest. The final blacklists for blood and airway myeloid and T cells appear in Table S8.

For both myeloid and T cells, we took all of the cells in the data set that we had annotated as each of these two cell types and used the drop-out score method described above to generate a list of putative, highly variable marker genes for each patient. Next, we generated a merged count matrix across all patients for a given cell type, which we normalized using the pooling method of Lun *et al* as described above (Lun et al., 2016). We then generated a log-normalized submatrix (log_2_(counts per million +1)) containing the union of the marker gene sets identified for each patient after removing genes on the airway and blood blacklists for the cell type-of-interest. Using the *PCA* function in the Python package *scikit-learn*, we decomposed this submatrix into its principal components. We used the 10 principal components with the largest eigenvalues as input to the scRNA-seq batch correction algorithm Harmony (Korsunsky et al., 2019). We made the function *HarmonyMatrix* aware of only the first 10 principal components and the patient identifiers for each cell. Finally, we computed a two-dimensional embedding using the Python implementation of the Uniform Manifold Approximation and Projection (UMAP) algorithm (McInnes and Healy, 2018) and the Pearson correlation matrix of the Harmony-corrected principal components. These embeddings appear in **Figures 2** and **4**.

For the differential expression analysis between blood and airway myeloid cells and between blood and airway T cells, we used the Mann-Whitney U-test approach described above. We removed genes on the blacklists described above for each cell type prior to subsampling, normalization, and statistical testing. We also restricted this analysis to protein-coding genes and removed all T cell receptor and immunoglobulin variable regions. We performed differential expression separately on each pair of matched airway and blood samples (there are 12 patient time points for which we have matched samples). Stringent criteria were used to select the differentially expressed genes displayed in the heatmaps in **Figures 2** and **4**. For the myeloid cell heatmap in **Figure 4**, a gene had to be differentially expressed with a fold-change of at least 4 in either direction and FDR<0.001 in at least 9 of the 12 matched sample pairs. For the T cell heatmap in **Figure 2**, we applied the first two criteria, but required them in only 6 of the 12 matched sample pairs. Results for all of the pairwise differential expression analyses comparing airway and blood T and myeloid cells can be found in Table S5 and Table S6, respectively.

### Analysis and visualization of Cytokine data

Cytokine expression data from early and late time points was log_10_ normalized and visualized as box plots overlaid with individual data points. The log_10_ normalized data was averaged across both the time points and used to generate heatmaps for cytokine expression across individual donor samples. Non-log transformed cytokine expression data from both time points was averaged for airway and plasma from each donor and used for paired-site analysis. Graphs were generated using the Python *matplotlib* and *seaborn* libraries (Hunter, 2007). All code for analysis of data and generation of figures will be hosted on GitHub.

### Lung tissue imaging analysis

Tissue segmentation was performed using inForm software on 10-30 representative fields (Version 2.3, PerkinElmer). Immune cell constituents within each tissue segment were defined by the DAPI nuclear counterstain to define the nucleus of each cell, with each associated membrane detected via presence of a specific stain (CD3, CD19, CD4, GzmB and/or CD163). Cell segmentation was adjusted as previously described to accurately locate all cells and minimize nuclear hypersegmentation and hyposegmentation (Weisberg et al., 2019). Cells were then phenotyped by training the phenotyping algorithm of inForm software, identifying: macrophage (CD163+ magenta cells), T cells (CD4^+^ cyan cells and CD8^+^ orange cells), B cells (CD19^+^ yellow cells). The cell segmentation data summary provided densities and numbers of each cell type in the lung tissue segments and the full cell segmentation data file provided the X and Y coordinates of each phenotyped cell.

### Statistical Analysis

Differences in mean between two sample groups were compared using Mann-Whitney U test, multiple group comparisons were done using ANOVA followed by Tukey’s HSD post-test and paired t-test for any paired data custom scripts based on Python *sciPy* library (Jones et al., 2001). *P*-values below 0.05 were considered as statistically significant. For all figures *** = p-value < 0.001, ** = p-value < 0.01, and * = p-value < 0.05.

## DATA AND CODE AVAILABILITY

The scRNA-seq data for each sample including count matrices, normalized counts, metadata, cell annotations, and UMAP embeddings are available on the COVID-19 Cell Atlas along with interactive visualizations (https://www.covid19cellatlas.org/index.patient.html). The scRNA-seq data analysis code is available at www.github.com/simslab/cluster_diffex2018.

## Supplementary Tables

**Table S1**. Clinical information for COVID-19 patients in this study.

**Table S2**. Assays performed on the samples from individual COVID-19 patients.

**Table S3**. Summary of sample details for scRNA-seq analysis.

**Table S4**. PCA loadings of markers for PC1 and PC2.

**Table S5**. Differential gene expression by T cells in airway versus blood for each sample by scRNA-seq.

**Table S6**. Differential gene expression by myeloid cells in airway versus blood for each sample by scRNA-seq.

**Table S7**. Deceased donors for control airway and COVID-19 lung autopsy samples

**Table S8**. Blacklisted genes for a given cell type for the scRNAseq analysis.

## Notes

### Competing Interest Statement

Jing Zhou, Matthew Steinle, and Sean Mackay have competing interests with IsoPlexis. The remaining authors declare no competing interests.

### Funding Statement

This work was supported by NIH grants AI128949 and AI06697 awarded to D.L.Farber and a Chan Zuckerberg Initiative COVID-19 grant awarded to D.L.Farber and P.A.Sims, and an R01AI093870 awarded to A.J.Y.

### Author Declarations

All human subjects research reported in this study was approved by the Columbia University institutional Review Board (IRB).

