## Supplementary figures and images for "Analysis of respiratory and systemic immune responses in COVID-19 reveals mechanisms of disease pathogenesis"

### Supplemental Figure1

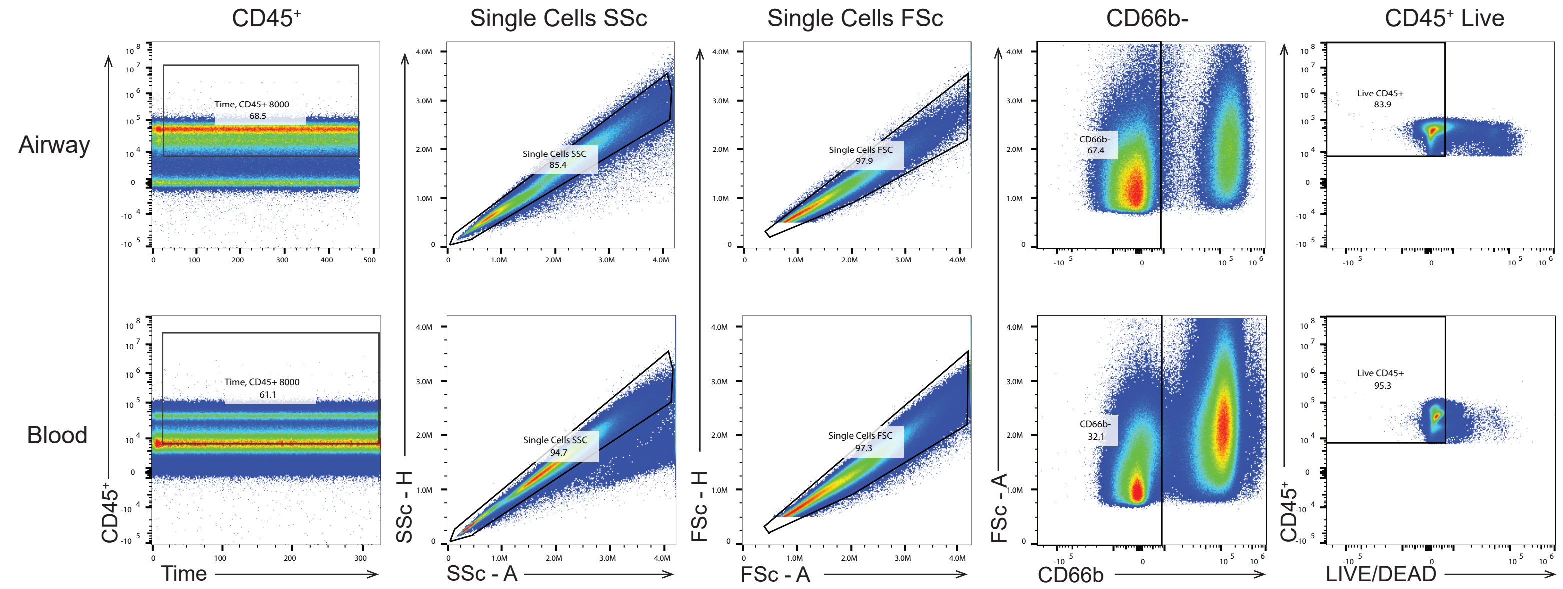

### Supplemental Figure2

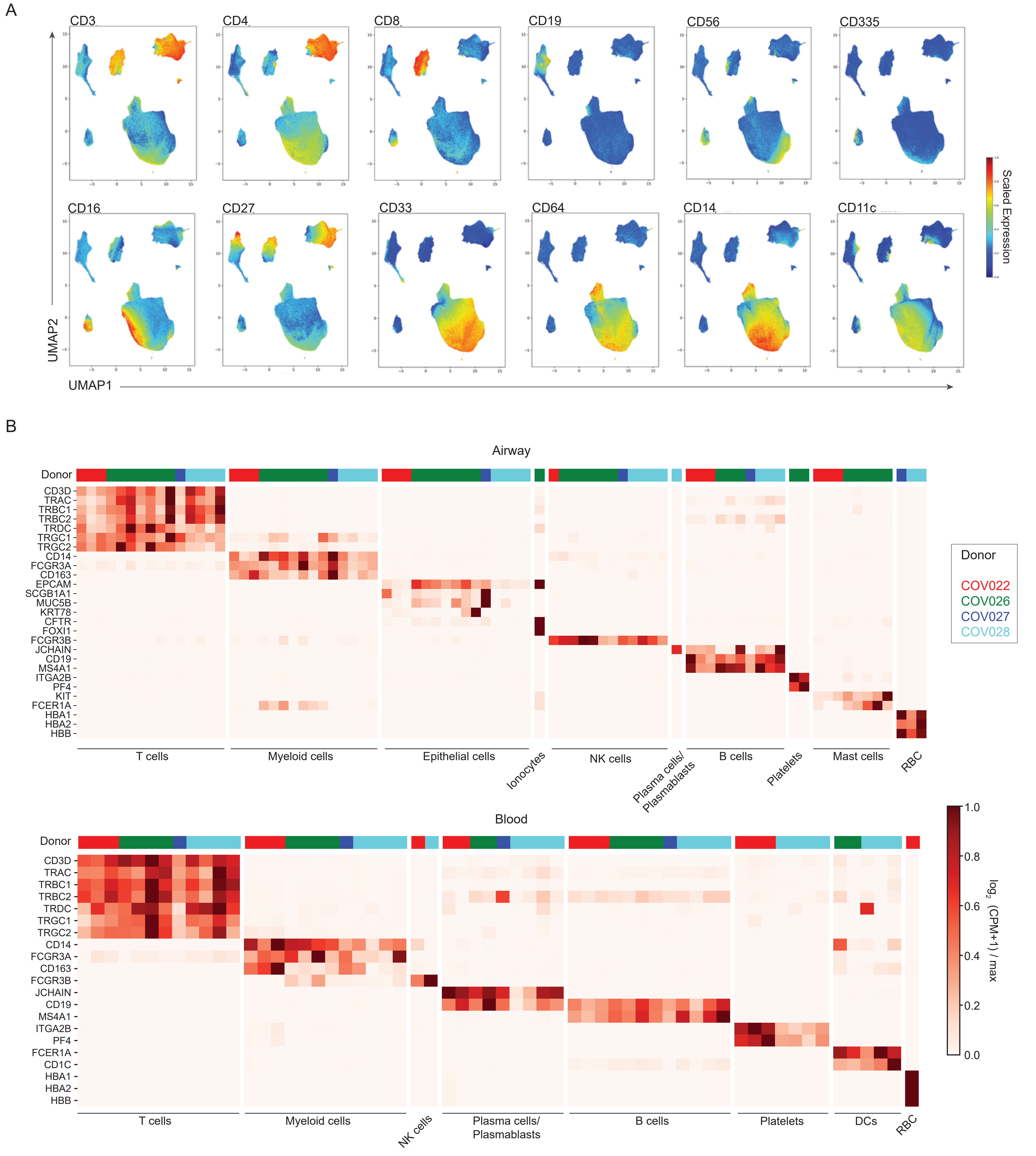

### Supplemental Figure3

Supplementary Figure 3.

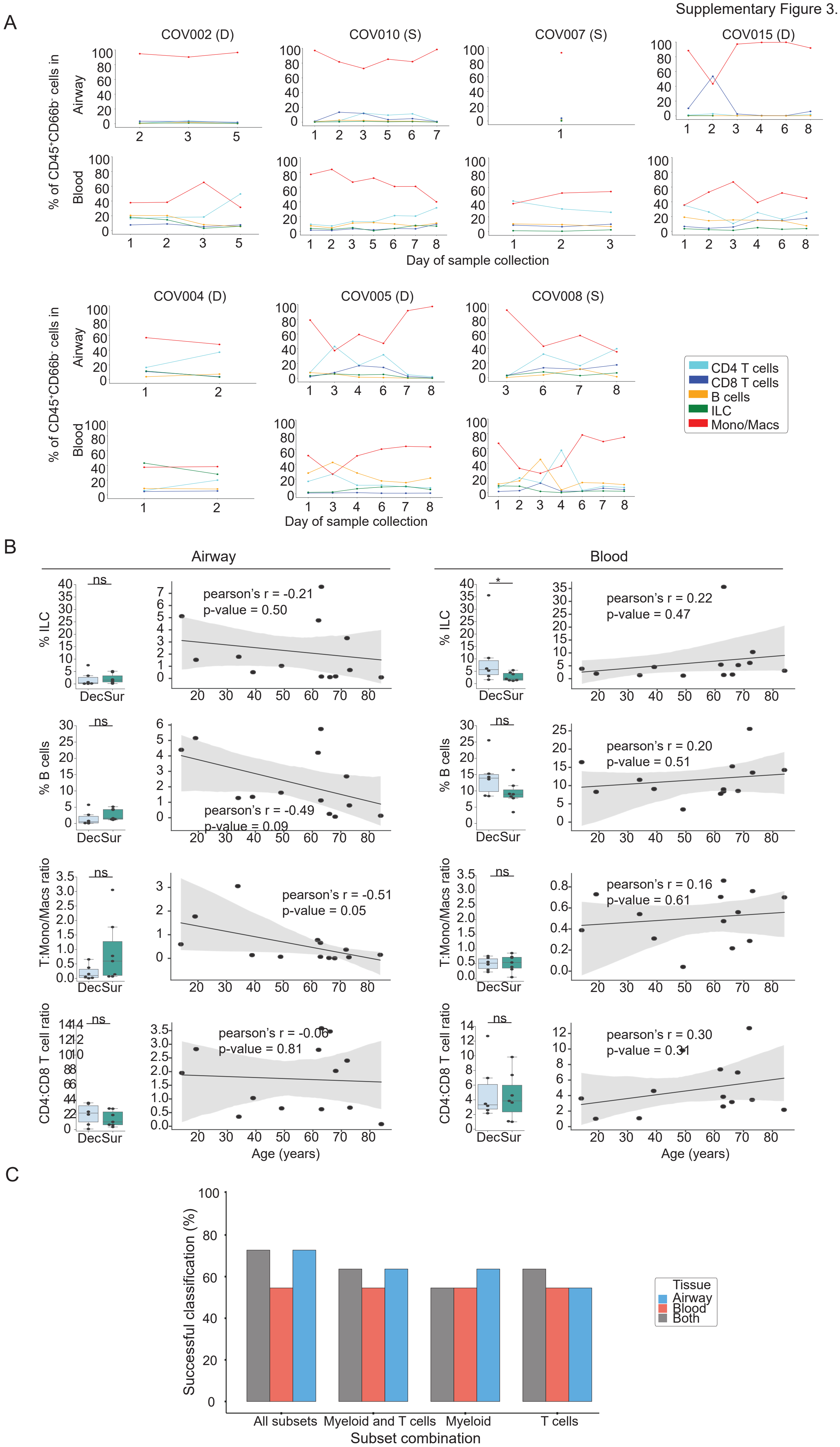

### Supplemental Figure4

Supplementary Figure 4.

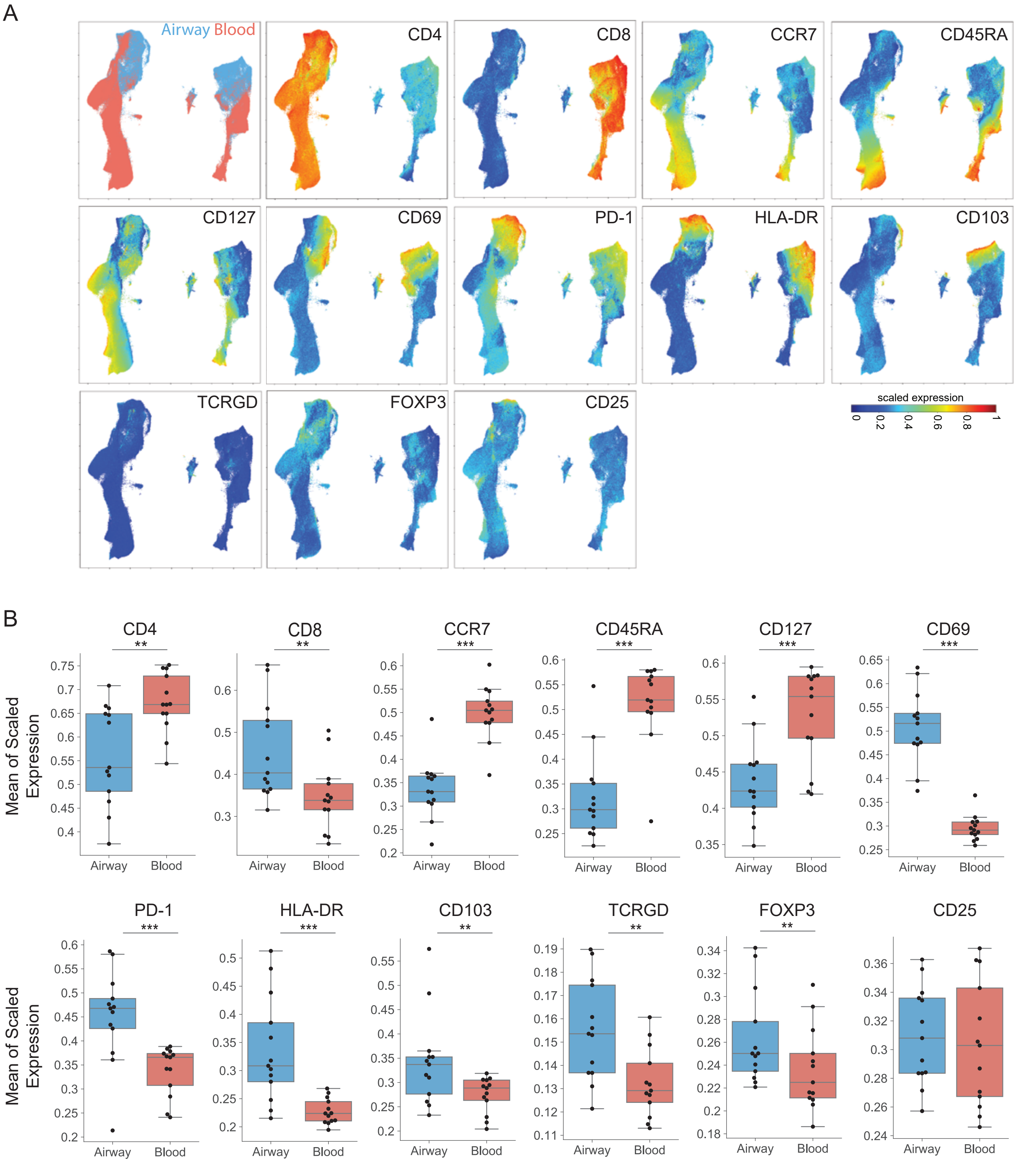

### Supplemental Figure5

Supplementary Figure 5.

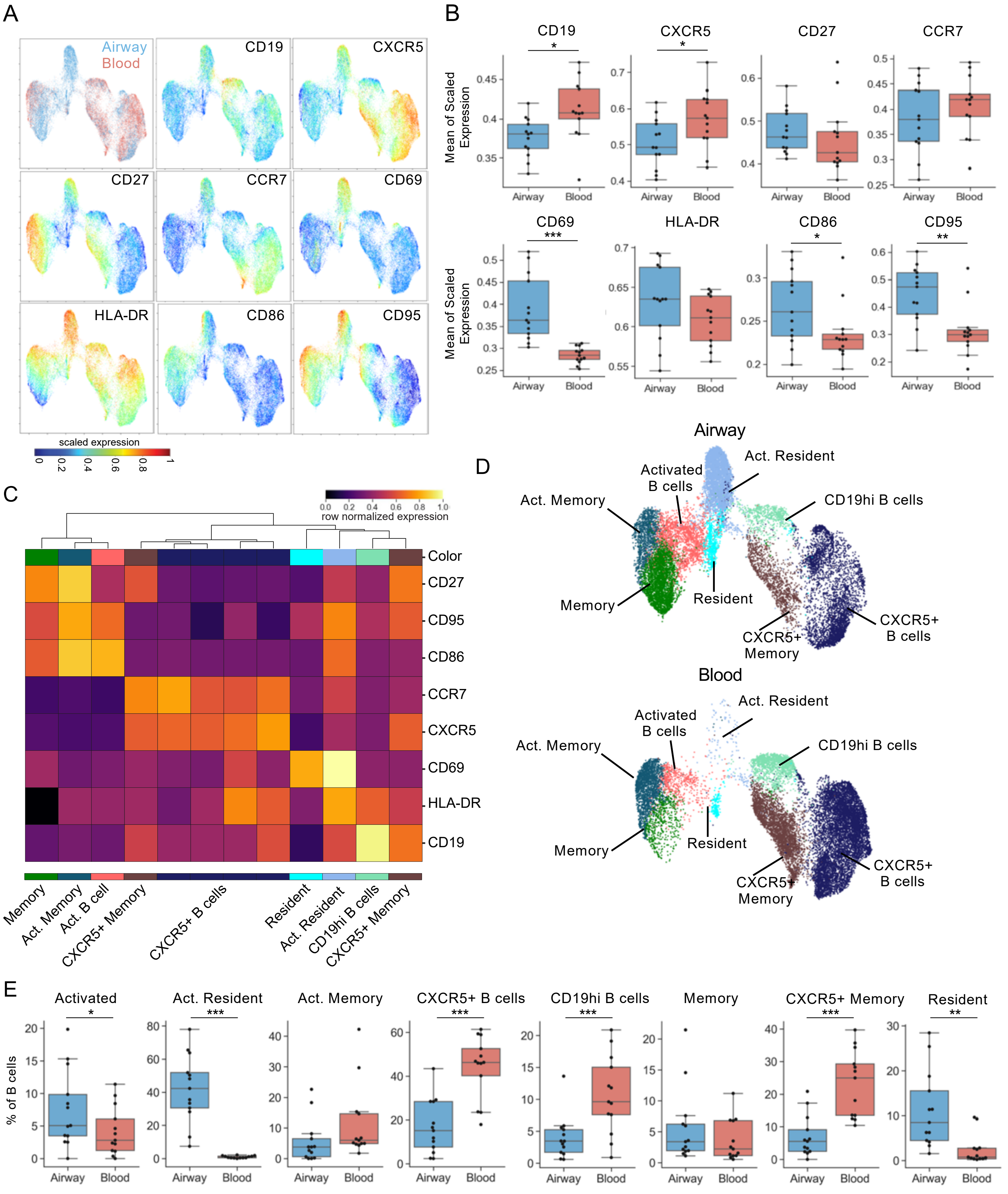

### Supplemental Figure6

Supplementary Figure 6.

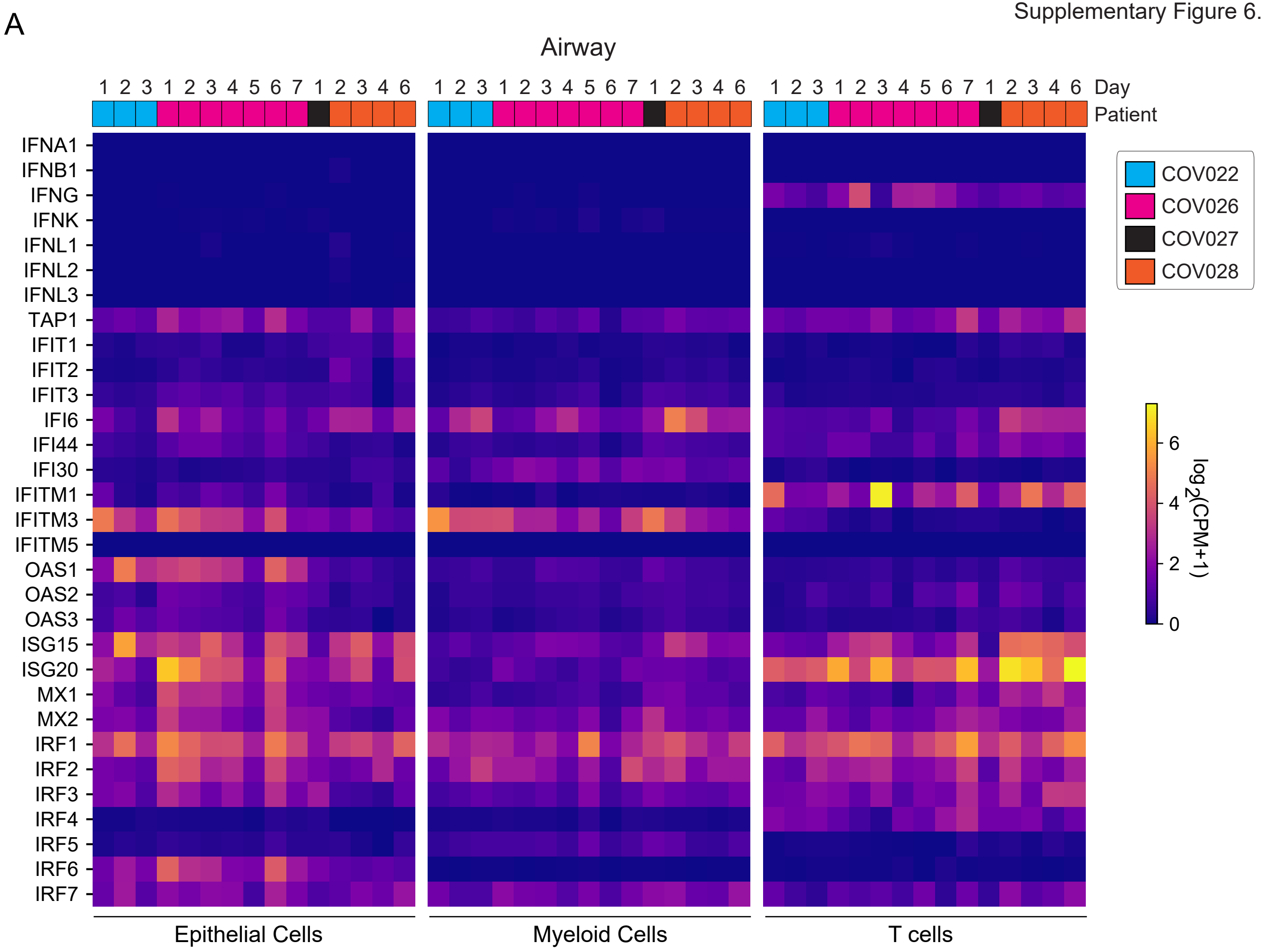

### Supplemental Figure7

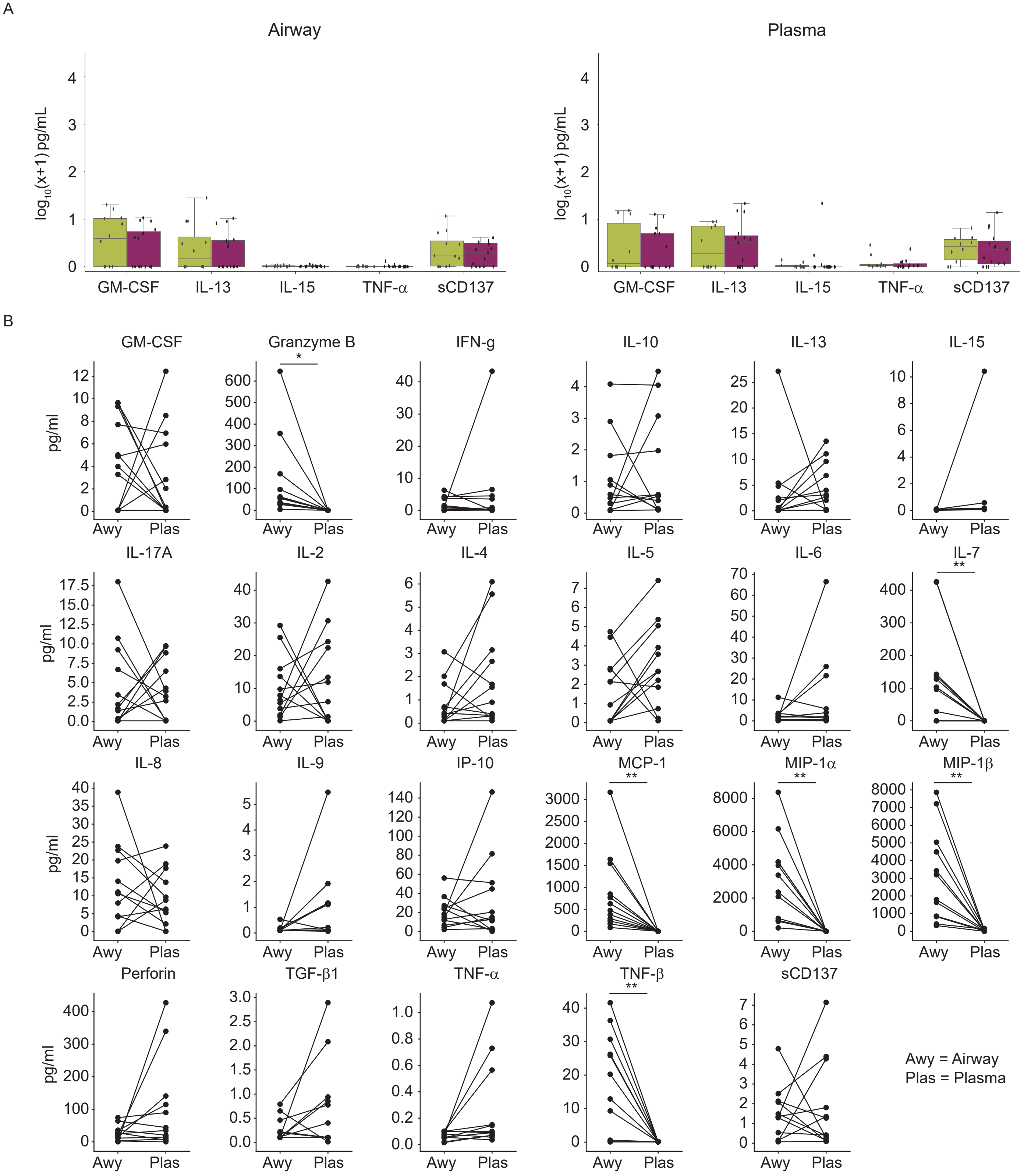
