## Supplemental Table1 for "Analysis of respiratory and systemic immune responses in COVID-19 reveals mechanisms of disease pathogenesis"

|  | **Deceased (n=8)** | **Survived (n=7)** | *P value* |
| --- | --- | --- | --- |
| **Clinical Characteristics** | | | |
| Age, years median (range) | 72.5 (63-84) | 39 (14- 63) | 0.0005 |
| Sex, male (%) | 5 (62.5%) | 4 (57.1%) |  |
| Body Mass Index, kg/m2, median (IQR) | 31.1 (29.3-35.0) | 36.3 (30.5-39.4) | 0.62 |
| SOFA, median (IQR)^a^ | 11.5 (10.3-14.8) | 12 (10.5-13.5) | 0.98 |
| Acute Respiratory Distress Syndrome (%) | 8 (100%) | 7 (100%) |  |
| SARS-CoV-2 PCR Positive | 8 (100%) | 7 (100%) |  |
| Days Post Symptom Onset, median (IQR)^b^ | 13.5 (8.8-16.5) | 12 (9-13) | 0.55 |
| Hospitalization, Days median (range) | 14.5 (1-36) | 43 (19-88) | 0.01 |
| **Race or Ethnic Group (%)^c^** | | | |
| Hispanic or Latino | 2 (25%) | 2 (28.6%) |  |
| Black or African American | 0 (0%) | 3 (42.9%) |  |
| White | 2 (25%) | 2 (28.6%) |  |
| Other or Unknown | 6 (75%) | 2 (28.6%) |  |
| **Co-Morbidities (%)** | | | |
| Hypertension | 6 (75%) | 1 (14.3%) |  |
| Diabetes | 3 (37.5%) | 2 (28.6%) |  |
| Chronic Cardiac Disease | 0 (0%) | 0 (0%) |  |
| Current or Former Smoker | 3 (37.5%) | 1 (14.3%) |  |
| COPD or ILD | 1 (12.5%) | 1 (14.3%) |  |
| Chronic Neurologic Diseases and Dementia | 1 (12.5%) | 0 (0%) |  |
| Asthma | 1 (12.5%) | 1 (14.3%) |  |
| **Laboratory Values, Median (IQR)^a^** | | | |
| Absolute Neutrophil Count, x10(3)/µL | 17.2 (12.7-23.2) | 15.1 (11.7-28.5) | 0.83 |
| Absolute Lymphocyte Count, x10(3)/µL | 0.95 (0.74-1.27) | 0.64 (0.52-1.05) | 0.52 |
| Absolute Monocyte Count, x10(3)/µL | 0.86 (0.63-1.26) | 0.77 (0.52-0.87) | 0.56 |
| D-Dimer, µg/mL^d^ | 7.5 (3.3-14.3) | 10.2 (2.8-17.9) | 0.99 |
| Ferritin, ng/mL | 1250 (636-2580) | 1767 (806-3929) | 0.69 |
| High Sensitivity C-reactive Protein, mg/L^d^ | 75 (38-157) | 94 (24-199) | 0.46 |
| Interleukin-6, pg/mL^d^ | 121.7 (86.9-315) | 64.5 (13.9-226.1) | 0.17 |
| Lactate Dehydrogenase, U/L | 646 (614-1129) | 598 (287-1052) | 0.44 |
| PaO2/FiO2 ratio | 122 (83-153) | 168 (119-180) | 0.40 |
| Abbreviations: SOFA - Sequential Organ Failure Assessment Score, COPD – Chronic Obstructive Pulmonary Disease, ILD – Interstitial Lung Disease, PaO2 – arterial partial pressure of oxygen, FiO2 – fraction of inspired oxygen | | | |
| a First available value within 48 hours of study enrollment | | | |
| b Days from onset of respiratory symptoms to study enrollment | | | |
| c Subjects included in all categories for which they identified | | | |
| d Values above upper limits of test entered as; D-Dimer (20mg/mL), C-reactive Protein (200mg/L), Interleukin-6 (315pg/mL) | | | |
| All statistical testing done with Mann-Whitney Testing | | | |

**Table S1.** Clinical information for COVID-19 patients in this study.
