## Supplemental Table2 for "Analysis of respiratory and systemic immune responses in COVID-19 reveals mechanisms of disease pathogenesis"

| Study ID | Age/sex | | Outcome | # of paired samples | Flow  Cytometry | Cytokines | scRNA-seq |
| --- | --- | --- | --- | --- | --- | --- | --- |
| COV002 | 73M | Deceased | | 4 | X | X |  |
| COV004 | 63M | Deceased | | 2 | X |  |  |
| COV005 | 72M | Deceased | | 7 | X | X |  |
| COV007 | 63F | Survived | | 4 | X | X |  |
| COV008 | 14F | Survived | | 10 | X | X |  |
| COV010 | 39M | Survived | | 6 | X | X |  |
| COV011 | 62M | Survived | | 7 | X | X |  |
| COV014 | 34F | Survived | | 7 | X | X |  |
| COV015 | 84M | Deceased | | 7 | X | X |  |
| COV022 | 49M | Survived | | 7 | X | X | X |
| COV024 | 68M | Deceased | | 7 | X | X |  |
| COV025 | 19M | Survived | | 8 | X | X |  |
| COV026 | 74F | Deceased | | 7 |  | X | X |
| COV027 | 82F | Deceased | | 1 |  |  | X |
| COV028 | 66F | Deceased | | 6 | X | X | X |

**Table S2.** Assays performed on the samples from individual COVID-19 patients.
