## Supplemental Table3 for "Analysis of respiratory and systemic immune responses in COVID-19 reveals mechanisms of disease pathogenesis"

| **Sample Name** | **Patient ID** | **Days Post-Intubation** | **Sample Source** | **Number of Cells** |
| --- | --- | --- | --- | --- |
| COV022A1 | COV022 | 1 | Airway Wash | 2,489 |
| COV022A2 | COV022 | 2 | Airway Wash | 9,852 |
| COV022A3 | COV022 | 3 | Airway Wash | 2,487 |
| COV022B1 | COV022 | 1 | Fresh PBMCs | 4,603 |
| COV022B2 | COV022 | 2 | Fresh PBMCs | 11,574 |
| COV022B3 | COV022 | 3 | Fresh PBMCs | 9,337 |
| COV026A1 | COV026 | 1 | Airway Wash | 4,651 |
| COV026A2 | COV026 | 3 | Airway Wash | 5,834 |
| COV026A3 | COV026 | 4 | Airway Wash | 4,147 |
| COV026A4 | COV026 | 5 | Airway Wash | 1,324 |
| COV026A5 | COV026 | 6 | Airway Wash | 2,834 |
| COV026A6 | COV026 | 7 | Airway Wash | 810 |
| COV026A7 | COV026 | 8 | Airway Wash | 2,065 |
| COV026B2 | COV026 | 3 | Frozen PBMCs | 5,608 |
| COV026B3 | COV026 | 4 | Frozen PBMCs | 5,080 |
| COV026B6 | COV026 | 7 | Frozen PBMCs | 4,846 |
| COV026B7 | COV026 | 8 | Frozen PBMCs | 4,755 |
| COV027A1 | COV027 | 1 | Airway Wash | 1,074 |
| COV027B1 | COV027 | 1 | Frozen PBMCs | 4,232 |
| COV028A2 | COV028 | 2 | Airway Wash | 7,025 |
| COV028A3 | COV028 | 3 | Airway Wash | 3,434 |
| COV028A4 | COV028 | 4 | Airway Wash | 2,598 |
| COV028A6 | COV028 | 7 | Airway Wash | 4,594 |
| COV028B2 | COV028 | 2 | Fresh PBMCs | 7,097 |
| COV028B3 | COV028 | 3 | Fresh PBMCs | 4,792 |
| COV028B4 | COV028 | 4 | Fresh PBMCs | 5,696 |
| COV028B6 | COV028 | 7 | Fresh PBMCs | 3,726 |

**Table S3.** Summary of sample details for scRNA-seq analysis.
