## Supplemental Table4 for "Analysis of respiratory and systemic immune responses in COVID-19 reveals mechanisms of disease pathogenesis"

| **Table S4.** PCA loadings of markers for PC1 and PC2. | | |
| --- | --- | --- |
| ­­ | **PC1** | **PC2** |
| CD3 | 0.09376378 | -0.00321972 |
| CD27 | 0.0541457 | -0.027022199 |
| CD8 | 0.042318087 | 0.022575237 |
| CD69 | 0.039878541 | 0.02799469 |
| CD4 | 0.037919562 | -0.009442391 |
| CD127 | 0.036891538 | 0.006012212 |
| CD45RA | 0.032480102 | -0.066146634 |
| CCR7 | 0.028651927 | -0.022445591 |
| CD28 | 0.028187697 | -0.000613115 |
| KLRG1 | 0.01210199 | 0.015968104 |
| CD103 | 0.009372095 | -0.003144 |
| PD-1 | 0.008226366 | 0.035477208 |
| CXCR5 | 0.007210185 | 0.00512664 |
| CD57 | 0.005966061 | -0.011104578 |
| CD19 | 0.004030206 | 0.01505254 |
| CD25 | 0.000343514 | 0.005715754 |
| FOXP3 | -0.00135812 | 0.030271536 |
| CD335 | -0.00285707 | 0.001084015 |
| CD56 | -0.004704473 | -0.017625799 |
| CD86 | -0.005843339 | 0.016512602 |
| CD16 | -0.013048941 | 0.001424049 |
| TCRGD | -0.01572467 | 0.02201 |
| CD95 | -0.020441971 | 0.002135385 |
| HLA-DR | -0.023708676 | 0.066405255 |
| CD64 | -0.069340649 | 0.018124553 |
| CD11C | -0.070580538 | 0.030887852 |
| CD163 | -0.072911997 | -0.03919522 |
| CD14 | -0.096348265 | -0.010123983 |
| CD33 | -0.123977753 | -0.032195659 |
