## Supplemental Table7 for "Analysis of respiratory and systemic immune responses in COVID-19 reveals mechanisms of disease pathogenesis"

| **Table S7.** Deceased donors for control airway and COVID-19 autopsy samples | | | | |
| --- | --- | --- | --- | --- |
| **Donor Number** | **Status** | **Sex** | **Age** | **Flow/IF** |
| D328 | Control | M | 52 | IF |
| D370 | Control | M | 52 | IF |
| D481 | Control | M | 29 | Flow |
| D484 | Control | M | 59 | Flow |
| D488 | Control | M | 55 | Flow |
| D489 | Control | F | 67 | Flow |
| D490 | Control | M | 26 | Flow |
| IMG001 | COVID-19 | M | 73 | IF |
| IMG002 | COVID-19 | F | 93 | IF |
| IMG003 | COVID-19 | M | 71 | IF |
| IMG004 | COVID-19 | F | 81 | IF |
| IMG005 | COVID-19 | M | 63 | IF |
